# Estimating the impact of disruptions due to COVID-19 on HIV transmission and control among men who have sex with men in China

**DOI:** 10.1101/2020.10.08.20209072

**Authors:** Ross. D. Booton, Gengfeng Fu, Louis MacGregor, Jianjun Li, Jason J. Ong, Joseph D. Tucker, Katy M.E. Turner, Weiming Tang, Peter Vickerman, Kate M. Mitchell

**Affiliations:** University of Bristol, Bristol, United Kingdom; MRC Centre for Global Infectious Disease Analysis, Department of Infectious Disease Epidemiology, Imperial College London, London, United Kingdom; Jiangsu provincial center for disease control and prevention, Nanjing, Jiangsu province, China; Social Entrepreneurship to Spur Health (SESH) Global, Guangzhou, China; Faculty of Infectious and Tropical Diseases, London School of Hygiene & Tropical Medicine, London, United Kingdom; Central Clinical School, Monash University, Melbourne, Australia; University of North Carolina Project-China, Guangzhou, China; University of North Carolina at Chapel Hill, Chapel Hill, United States

**Keywords:** COVID-19 pandemic, men who have sex with men, modeling, HIV transmission, key and vulnerable populations, People’s Republic of China

## Abstract

**Introduction:** The COVID-19 pandemic is impacting HIV care globally, with gaps in HIV treatment expected to increase HIV transmission and HIV-related mortality. We estimated how COVID-19-related disruptions could impact HIV transmission and mortality among men who have sex with men (MSM) in four cities in China.

**Methods:** Regional data from China indicated that the number of MSM undergoing facility-based HIV testing reduced by 59% during the COVID-19 pandemic, alongside reductions in ART initiation (34%), numbers of sexual partners (62%) and consistency of condom use (25%). A deterministic mathematical model of HIV transmission and treatment among MSM in China was used to estimate the impact of these disruptions on the number of new HIV infections and HIV-related deaths. Disruption scenarios were assessed for their individual and combined impact over 1 and 5 years for a 3-, 4- or 6-month disruption period.

**Results:** Our China model predicted that new HIV infections and HIV-related deaths would be increased most by disruptions to viral suppression, with 25% reductions for a 3-month period increasing HIV infections by 5-14% over 1 year and deaths by 7-12%. Observed reductions in condom use increased HIV infections by 5-14% but had minimal impact (<1%) on deaths. Smaller impacts on infections and deaths (<3%) were seen for disruptions to facility testing and ART initiation, but reduced partner numbers resulted in 11-23% fewer infections and 0.4-1.0% fewer deaths. Longer disruption periods of 4 and 6 months amplified the impact of combined disruption scenarios. When all realistic disruptions were modelled simultaneously, an overall decrease in new HIV infections was always predicted over one year (3-17%), but not over 5 years (1% increase-4% decrease), while deaths mostly increased over one year (1-2%) and 5 years (1.2 increase – 0.3 decrease).

**Conclusions:** The overall impact of COVID-19 on new HIV infections and HIV-related deaths is dependent on the nature, scale and length of the various disruptions. Resources should be directed to ensuring levels of viral suppression and condom use are maintained to mitigate any adverse effects of COVID-19 related disruption on HIV transmission and control among MSM in China.

## Introduction

Globally, 37.9 million people are living with HIV (PLWH) [1], with men who have sex with men (MSM) disproportionally affected [2]. In China, a recent systematic review indicated an HIV prevalence of 5.7% (95% CI:5.4-6.1%) among MSM, with increasing HIV prevalence in this group over time (2001-2018) [3]. Efforts to manage the HIV epidemic in China have been made increasingly difficult by the emergent COVID-19 pandemic [4,5], having significant potential to affect the HIV care continuum and patterns of sexual risk behaviour in numerous settings worldwide [6–10]. Close examination of this syndemic is a key issue for global public health [11].

Among PLWH in China - who already face high levels of HIV stigma, psychological distress, and suboptimal adherence to antiretroviral therapy (ART) [4] - the COVID-19 pandemic has presented further barriers to HIV control [5]. Quarantine and social distancing reduce access to routine HIV testing which reduces the identification and treatment of new HIV infections [12]. Timely linkage to HIV services and initiation of ART have been affected during the COVID-19 pandemic, with many hospitals designated for treatment of COVID-19 suspending taking on new patients with HIV [4]. The COVID-19 pandemic has hindered ART, due to hospital visits being restricted from city lockdowns/traffic controls [4]. In February 2020, a survey in China found 32.6% of PLWH were at risk of ART discontinuation and about half (48.6%) did not know where to get ART in the near future [13]. These gaps in HIV treatment could lead to increased HIV-related deaths and higher risk of HIV transmission. Conversely, MSM in other countries have reported having fewer sexual partners during periods of COVID-19-related lockdown, which may temporarily reduce HIV transmission [7].

Mathematical modelling can be used to capture the complexity of these changes and estimate the impacts of COVID-19 on HIV epidemiology. One modelling study of PLWH in Africa projected that a 6-month interruption in ART supply across 50% of the population on treatment could lead to a 60% increase in HIV-related deaths over a 1-year period [14]. Another modelling study on low-and-middle-income countries (LMIC) projected that HIV-related deaths would increase by 10% over the next five years, with the greatest impact on mortality estimated to be from ART interruptions [15]. However, neither of these studies used observed COVID-19 impact data to inform their modelled disruptions, which is essential for obtaining reliable projections for the true scale of COVID-19 disruptions.

To our knowledge no COVID-19 impact modelling has been published focussing on key populations, who are the main groups affected by HIV [16]. In addition, no model projections have to date incorporated observed data from the COVID-19 disruption. In this study, we addressed this by collating data on the impact of COVID-19 and resulting lockdown measures on HIV testing and treatment among PLWH, sexual risk behaviour and condom use among MSM in China. We used a deterministic model of HIV transmission and treatment among MSM in China to estimate the impact of these disruptions on new HIV infections and HIV-related deaths. We identified where HIV prevention and treatment efforts should be focussed to help mitigate potential adverse effects of COVID-19.

## Methods

### Observed disruptions due to COVID-19 in China

Estimates of changes in HIV testing (among MSM), treatment initiation and viral load suppression (among all PLWH) came from surveillance data from Jiangsu province (HIV testing/clinics), from the first quarter of 2019 and 2020 [17]. Estimates of changes in number of sexual partners and consistency of condom use came from an online survey conducted among MSM (N=731) across 31 provinces in China between 18/05/2020-02/06/2020, during the COVID-19 pandemic. From these data, we estimated the following percentage changes due to the disruption caused by COVID-19, compared to the pre-COVID period (before 01/01/2020):

I. The number of MSM undergoing facility-based HIV testing in the first quarter of 2019 was 6436 compared to 2641 in 2020, a reduction of 59% (95% CI:58-60%).
II. The number of PLWH initiating ART in the first quarter of 2019 was 315 compared to 208 in 2020, a reduction of 34% (95% CI:29-39%).
III. There was no change in viral suppression (VS) among PLWH. 95.3% (940/986; 95% CI:93.8-96.6%) of viral load tests showed VS in the first quarter of 2019, with similar numbers in 2020 (96.0%, 928/967; 95% CI:94.5-97.1%). The proportion of diagnosed PLWH who had a viral load test was similar in both years: 4.7% in the first quarter of 2019 and 4.3% in the first quarter of 2020. Note most viral load tests in this region are conducted in the 3^rd^/4^th^ quarter.
IV. 62% (313/506) of MSM reported reduced numbers of sexual partners compared to the pre-pandemic period (among those who had male partners in the last six months).
V. 25% (126/506) of MSM used fewer condoms with their partners compared to the pre-pandemic period.

### Mathematical model

We used a model of HIV testing/transmission/treatment among MSM in China which was previously developed to evaluate the long-term impact of an HIV self-testing intervention in four cities (Guangzhou/Shenzhen/Jinan/Qingdao) [18]. All individuals within the model are categorised by infection status, risk (≤/>two male anal sex partners in the last three months), anal sex role (always insertive/versatile/always receptive), infection stage (acute/CD4>500/351-500/200-350/<200 cells/µl), and diagnosed/ART status. Uninfected MSM enter the population upon sexual debut and leave through migration or non-HIV/HIV-related death. Those not on ART move into more advanced stages of infection, while those on ART do not; their mortality is modelled as a function of infection stage at ART initiation (Fig.S2).

Both facility-based and self-testing are modelled, and MSM are distinguished by whether or not they have previously tested. Those on ART who drop out re-enter the diagnosed compartments. HIV transmission occurs via anal sex between MSM at a rate which depends on HIV disease stage/ART coverage/VS/total partners/total sex acts/sexual role/condom efficacy and use. The model was calibrated to MSM city-level HIV epidemics, parameterised using demographic/behaviour data from CDC/trials, calibrated to local city/province/national-level estimates (“fitting metrics”) of HIV prevalence/ART coverage/diagnosis/incidence/population size [18]. The model was solved in R.3.5.1. Model/schematics (Figs.S2-3), parameters/fitting metrics (Table S6) and fitting metrics (Fig.S1) are given in the supplement.

### Base case scenario (no COVID-19)

The model was run for five years until 01/01/2025 using the fitted model parameters, with all parameters constant at their 2019 values from 01/01/2020 onwards. These base case runs predicted the non-COVID-19 trajectory of HIV prevalence and care for each city.

### COVID-19-related disruption scenarios

Disruption scenarios were implemented from 01/01/2020 and run for 3 months, after which all parameters were reset to their original pre-COVID-19 values. Comparisons of these scenarios with the base case were made over one (01/01/2021) and five years (01/01/2025).

The following ***observed*** disruption scenarios were based on the observed disruptions – reductions in:0

A. facility-based HIV testing (59%)
B. ART initiation (34%)
C. number of sexual partnerships (31–62%)
D. condom use (12.5–25%)

Although data from Jiangsu province suggested no disruption to VS, disruptions in ART provision have been reported to the WHO [19]. We explored an additional ***hypothetical*** scenario where VS was reduced by 10% (consistent with reductions in ART access among MSM in United States [7]) and 25% (consistent with disruptions to ART uptake reported among PLWH in China [4]):

E. Reduction in VS of 10/25%

The data on sexual partnerships and condom use estimated the proportion of MSM having fewer partnerships/using condoms less frequently (not of their overall reductions). We sampled partnership and condom use parameters from uniform distributions with bounds of half-reported to full-reported reductions. Reductions in condom use were modelled as a reduction in the proportion of sex acts in which a condom is used. Reductions in HIV testing and ART initiations were modelled as reductions in the facility-based HIV testing rate and ART initiation rate, and reductions in partner numbers were modelled as reductions in numbers of partners per year (applied across all risk groups). VS reductions were modelled as increases in infectiousness and HIV-related mortality among those on ART, assuming a proportion (10%/25%) of virally suppressed MSM stop taking ART, having the same infectiousness and HIV-related mortality as individuals not on ART. No reduction in HIV self-testing rates were modelled, in line with local observations in Jiangsu.

We assessed the impact of each disruption (A,B,C,D,E) separately, and also assessed the combined impact of these disruptions occurring simultaneously (±scenario-E). The impacts of disruptions lasting 3, 4 and 6 months were assessed.

The outcome measures used to assess the disruption caused by COVID-19 were total and relative percentage change in new infections and HIV-related deaths, compared to the base case, non-COVID-19 scenario. These impacts were evaluated over 1-year (01/01/2020-01/01/2021) and 5-years (01/01/2020-01/01/2025). All impact measures were expressed as median values and 95% credible intervals (95% CrI), across the 100 selected parameter sets in each city, and across the 400 parameter sets from all cities.

We analysed the sensitivity of these scenarios (A,B,C,D,E) to different magnitudes of disruption (0,25,50,75,100%) over 1 and 5 years with a 3-month disruption. We plot the % change in new HIV infections and HIV-related deaths as a function of each individual disruption parameter.

## Results

The percentage change in impact measures (new HIV infections and HIV-related deaths) did not vary between each city, with greater within-city variation across the scenarios. Therefore, all results are presented as the overall impact across four cities (Table S1, Guangzhou/Shenzhen/Jinan/Qingdao), with results for each city in the supplement (Tables S2-5).

### Single and combined 3-month disruptions

Realistic disruptions to facility-based HIV testing, ART initiation and condom use were each estimated to lead to an increase in new HIV infections among MSM (Fig.1a, Tables S1-S5). Disruptions to condom use (scenario-D) lasting 3 months were predicted to lead to the largest overall relative increase in HIV infections, of 7.8% (95% CrI:4.5-13.8%) over one year (Fig.1a), with relative increases of 2.3% (1.7-2.9%) and 1.7% (1.2-2.4%) predicted over 1 year for realistic 3-month disruptions to facility-based HIV testing (scenario-A) and ART initiations (scenario-B), respectively. Reductions in numbers of sexual partners (scenario-C) were predicted to reduce HIV infections, by a median 16.2% (11.1-23.2%) over one year among MSM following a 3-month disruption.

**Figure 1:**
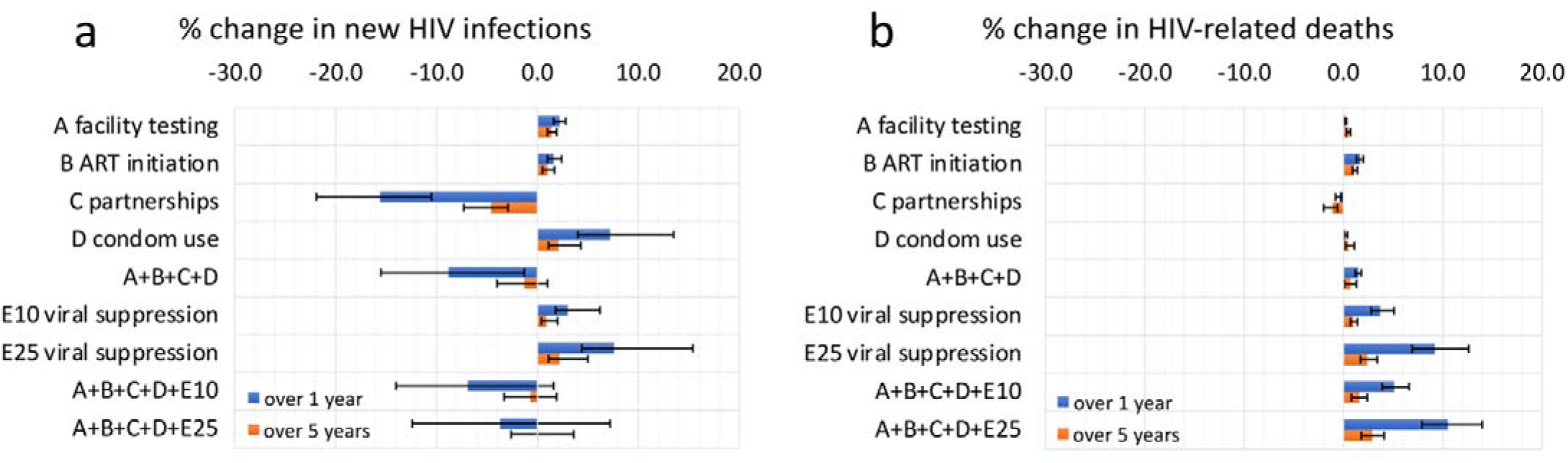
The percentage change in (a) new HIV infections and (b) HIV-related deaths under disruption scenarios evaluated over a 1- and 5-year time horizon (blue and orange respectively) in four cities in China. Bars indicate median values, while error bars show the 95% credible intervals for each scenario and time horizon. Scenarios are as follows: A) Reduction in facility-based HIV testing (59%), B) Reduction in ART initiation (34%), C) Reduction in number of sexual partnerships (31 – 62%), D) reduction in condom use (12.5 – 25%) E10) Reduction in viral suppression of 10%, E25) Reduction in viral suppression of 10%.

A hypothetical 10%/25% reduction in VS (E10/E25) would lead to increased numbers of HIV infections. 25% reductions in VS increased new HIV infections by 7.4% (4.7-14.0%) over one year given a 3-month disruption.

The effect of each disruption scenario on the relative percentage change in HIV infections was always smaller over five years than after one year (but not the absolute difference in HIV infections, which generally increased over 5 years), with a more rapid decrease in effect over 5 years seen for disruptions to partnership numbers, condom use and VS (Fig.1a).

When all of the observed disruptions (A+B+C+D) were modelled simultaneously, a decrease in new HIV infections was always predicted over one year (median 8.7%, (2.8%-17.2%)), but over 5 years this impact reduced to a 1.6% (−0.6%-4.3%) decrease in new HIV infections due to the disruptions to HIV testing (increase 1.7%, 1.2%-2.4%) and ART initiation (increase 1.1%, 0.7-1.8%) having a longer-lasting effect on ART outcomes, with ART taking longer post-disruption to return to pre-disruption levels.

HIV-related deaths were also predicted to increase following disruptions to HIV testing/ART initiations/condom use/VS (scenarios-A,B,D,E), and decrease following disruptions to partnerships (scenario-C) (Fig.1b, Tables S1-S5). Small impacts (<1%) on HIV-related deaths were predicted over one year for 3-month disruptions to HIV testing, partner numbers or condom use. Larger increases in HIV-related deaths were predicted to occur following 3-month disruptions to ART initiations – a 1.8% (1.5-2.0%) increase over one year–and, especially, VS – a 10.1% (7.6-12.7%) increase over one year following a 25% VS reduction. The observed disruptions together (A+B+C+D) resulted in 1.5% (1.1-1.8%) more HIV-related deaths over one year and 0.6% (−0.3-1.2%) over five years.

Including reductions of 10%/25% in levels of VS alongside the other, observed scenarios (A+B+C+D+E10, A+B+C+D+E25) always led to an increase in new HIV infections, and, particularly, an increase in HIV-related deaths, over a 1- or 5-year time horizon, compared to the observed scenarios (A+B+C+D) alone (Figs.1a,b).

### Sensitivity in disruption duration

When comparing the impacts of two combined disruption scenarios (A+B+C+D and A+B+C+D+E25) for disruptions lasting 3/4/6 months, we found impacts on both new HIV infections and HIV-related deaths were approximately linear, with a 4-month disruption leading to around 34% greater impact than a 3-month disruption, and a 6-month disruption around two times the impact of a 3-month disruption, over both the 1- and 5-year time horizons (Fig.2).

**Figure 2:**
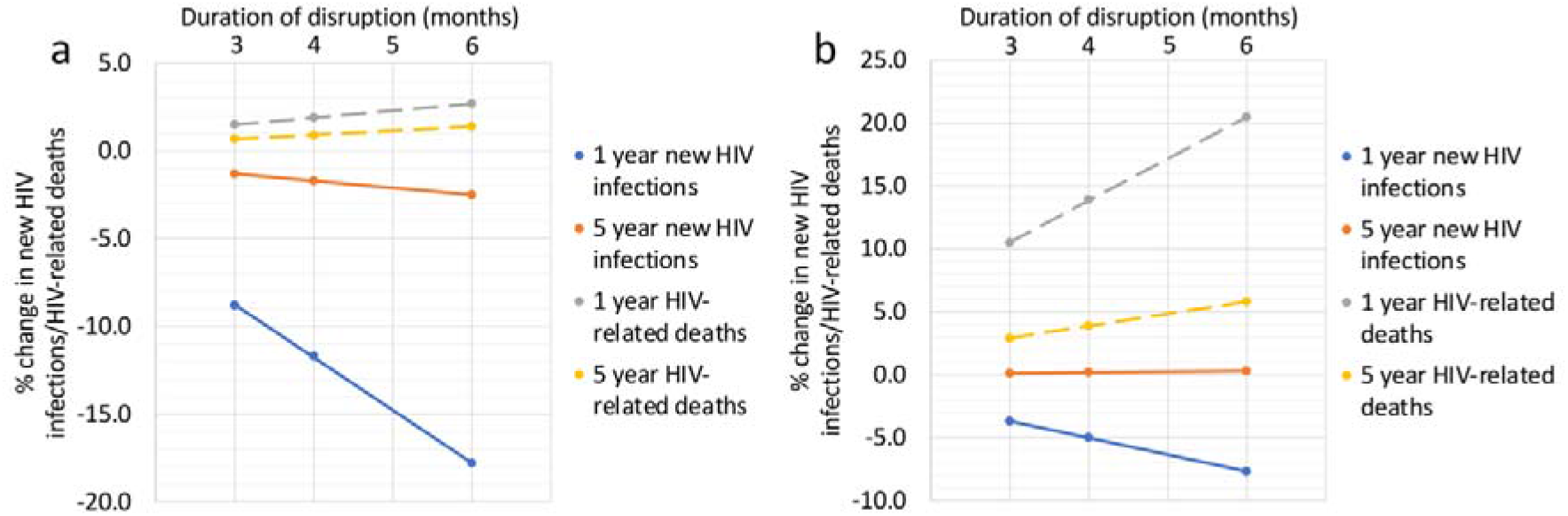
The percentage change in new HIV infections and HIV-related deaths for scenarios (a) A+B+C+D and (b) A+B+C+D+E25 for varying disruption periods (3, 4 and 6 months) and time horizons (1- and 5-year) in four cities in China. Dots indicate median values.

### Impact in different cities

The absolute numbers of predicted additional/prevented infections and deaths varied for each city (Tables S2-S5), related to differing MSM population sizes, but the percentage changes in infections and deaths did not vary substantially between cities (Fig.3, Tables S2-S5). For example, for scenario A+B+C+D, the overall predicted reduction in new HIV infections over one year varied from 8.7% (−2.8-17.2%) in Qingdao to 9.1% (2.5-15.0%) in Jinan, with far greater within-city than between-city uncertainty (Fig.3).

**Figure 3:**
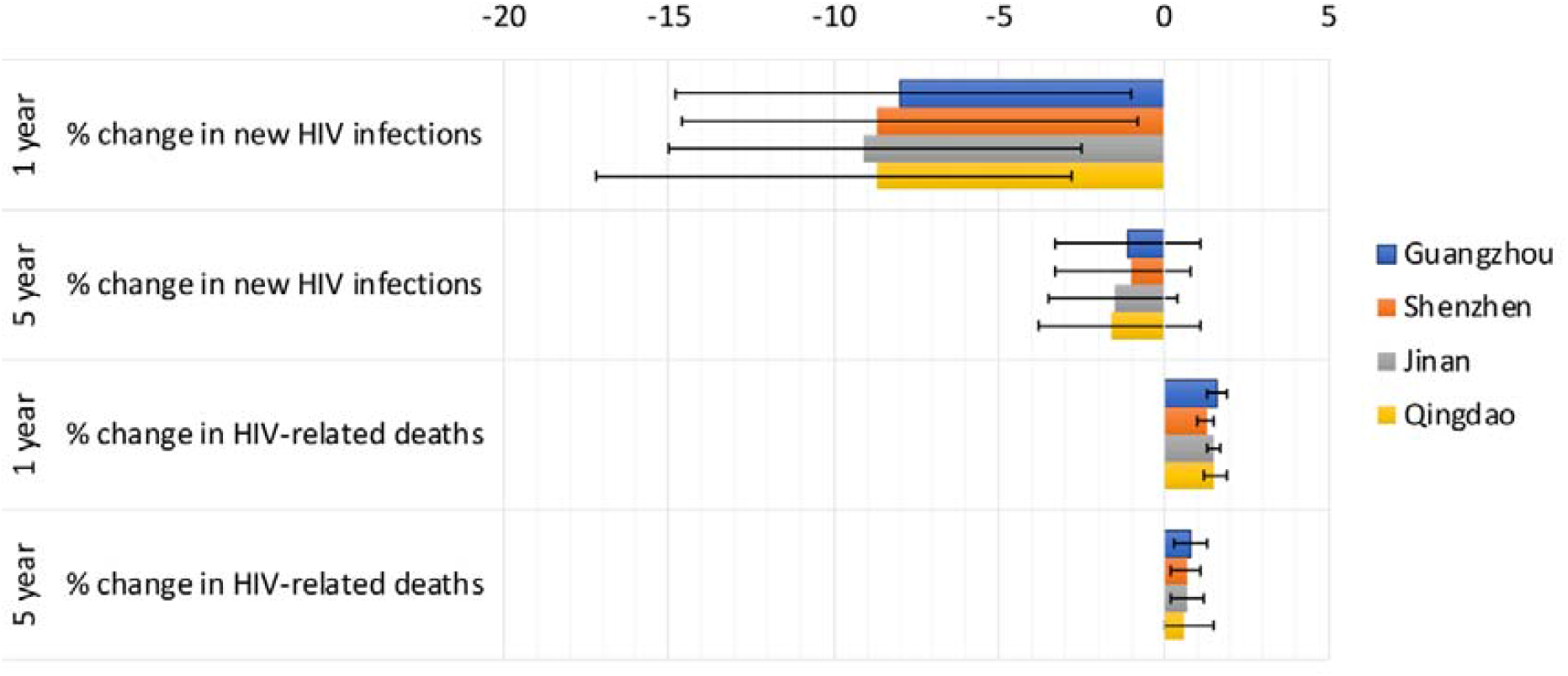
The percentage change in new HIV infections and HIV-related deaths for scenario A+B+C+D for different cities (Guangzhou, Shenzhen, Jinan and Qingdao), and time horizons (1- and 5-year). Bars indicate median values and error bars show the 95% credible intervals.

Over 5 years, realistic 3-month disruptions to HIV testing, ART initiations, sexual risk behaviour and condom use (scenario A+B+C+D) would lead to on average 3 fewer new HIV infections but 1 additional HIV-related death among MSM in Jinan and Qingdao, 6 fewer infections and 3 additional deaths in Guangzhou, and to 18 fewer HIV infections but 9 additional HIV-related deaths among MSM in Shenzhen.

The combined hypothetical scenario (A+B+C+D+E) lasting for 3 months, over 5 years, would lead to an average 3 additional new HIV infections in Guangzhou, 7 in Shenzhen but no change in Jinan and Qingdao, with 11 additional HIV-related deaths in Guangzhou, 33 in Shenzhen and 3 in Jinan and Qingdao.

### Sensitivity in disruption magnitude

Over 1 or 5 years, with a 3-month disruption, the relationship between the magnitude of the disruption (0,25,50,75,100%) and the projected impact was always linear (Fig.S4-5, Table S7-8), with higher values of % disruption leading to increases in new HIV infections and HIV-related deaths for four disruption parameters (facility testing/ART initiation/condom use/VS) but not for partnership disruption, (fewer infections and deaths).

For a 3-month disruption evaluated over a 1-year, theoretical disruption scenarios A-E50 (affecting 50% of MSM) increased new HIV-infections by 1.8%, 2.4%, −16.7% (decrease), 15.8% and 11.3% respectively, and increased HIV-related deaths by 0.1%, 2.5%, −0.4% (decrease), 0.5% and 18.2% respectively (Fig.S4, Table S7). If disruption scenarios affected 100% of MSM (A-E100), then new HIV-infections were projected to increase to 3.7%, 4.9%, −30.2% (decrease for 95% affecting MSM – the model requires >0 partnerships), 38.7% and 29.3% respectively. Scenarios A-E100 would also cause HIV related deaths to increase to 0.3%, 5.3%, −0.8% (decrease – for 95% disruption), 1% and 35.7% respectively.

## Discussion

Available data in this analysis suggests that the COVID-19 pandemic and measures undertaken against it have resulted in reduced rates of HIV testing and treatment among MSM in this region of China but have not so far had an impact on VS rates. Survey data also suggested MSM in China had fewer partners and used condoms less often during the COVID-19 pandemic. Using these data in our modelling analysis, simulating realistic 3-month disruptions to HIV testing, ART initiation, condom use and partner numbers, we found fewer new HIV infections are projected to occur among MSM in China over 2020 (9% fewer) than would have occurred in the absence of the COVID-19 pandemic, with a smaller decrease (2%) seen over 5 years. This decrease was largely due to reductions in sexual partner numbers counteracting a reduction in ART initiations and condom use. Our models do suggest these disruptions will lead to small increases in HIV-related deaths (2% over 1 year and 1% over 5 years). When we also evaluated potential reductions in VS of 25% alongside these observed disruptions, we predicted a 4% decrease in new HIV infections and 10% increase in HIV-related deaths over one year. When evaluating the effects of each individual disruption separately, new HIV infections were most adversely affected by disruptions in condom use and VS, and HIV-related deaths by reductions in VS. Therefore, our results suggest HIV prevention and treatment efforts should focus on maintaining use of condoms and VS among MSM in China to mitigate short- and long-term adverse effects of the COVID-19 disruption.

Although disruptions to VS had consequences for HIV incidence/mortality, data from Jiangsu [17] indicated no change in VS among PLWH due to COVID-19 disruption. Other surveys among PLWH in China indicate disruptions to ART access [4] which could lead to reductions in VS. It is critically important to quantify such reductions in treatment access and VS among MSM in China, and we suggest that future surveys should focus their efforts on determining the true scale of the disruption to VS, which is likely to be delayed and could occur after observed treatment disruptions.

The length of disruption is also critical in determining the longer-term impacts of COVID-19. Throughout our main analysis we used a disruption length of 3 months, with sensitivity analyses (with longer 4/6-month disruptions) demonstrating a linear relationship between the duration of disruption and the change in both new HIV infections and HIV-related deaths, with a doubling in disruption duration leading to a doubling in the impact. When assessing the combined impact of the observed disruptions, the direction of the linear relationship was different for HIV-related deaths (positive – longer duration gave more deaths) and new HIV infections (negative – longer duration gave fewer new infections).

The absolute numbers of new HIV infections and HIV-related deaths varied between the four cities in China modelled, due to the different population sizes, epidemiology and care cascade in each city (reflected in data used for calibration [18]). However, the percentage change in impact measures did not vary between each city, with much greater within-city variation. This result is surprising, considering each city has different future projected HIV prevalence (5.0 – 12.2% in 2036, based on data specific to these cities [20] in Booton et al. [18]) and are in two different provinces (Guangdong/Shandong). Therefore, the impact predicted in this study is likely to be applicable to MSM within any city/region in China.

We may compare our results to other modelling studies predicting the potential impact of COVID-19 related disruption on HIV prevention and treatment. Jewell et al. [14] used multiple African models of 3 month disruptions affecting 50% from 01/04/2020, reporting increases in HIV incidence of <1% from the suspension of HIV testing (*compared to our prediction of* 1-2% for *50% reductions* in facility testing, scenario-A50, all scenarios in Table S7), <2% from no new ART initiations (2-4%, scenario-B50), 2-9% from the interruption of condom availability (12-33%, scenario-D50), 4-89% from ART interruption (9-31% for 50% reduction in VS, scenario-E50). Suspension of testing for 50% increased deaths by <1% (<1%, scenario-A50), ART initiation <2% (2-3%, scenario-B50) and condom availability 0% (0-1%, scenario-D50) with ART interruption causing an increase of 17-62% (14-25%, scenario-E50). Our results align well with these estimates, considering the different methodology/definitions of disruption/population (all adults/children, compared to solely MSM) and underlying models and data (different settings/treatment/condom use). This is a major strength of our study, the use of early data from China, to estimate data-driven (rather than theoretical) magnitudes of COVID-19-related disruptions. Another major strength of this study was performing our analysis on four separate cities from two distinct regions within China. In addition, our analysis involves various scenarios and the effects of combining these enables us to better understand the relative impact of different disruptions.

Generalising our results to LMIC should be done with consideration of the differences between China and the respective country. Early estimates of COVID-19 impacts on testing/treatment/sexual risk behaviour were available as China was the first centre of the COVID-19 pandemic, and as more data becomes available, other LMIC countries may report different survey data. Our modelling analysis was able to highlight which of these disruptions are likely to have the biggest negative impacts on HIV incidence and deaths, indicating disruptions which should be prioritised for monitoring and mitigation in other countries.

Our analysis has some limitations which should be acknowledged. Not all of the disruption estimates were MSM specific, and MSM may have had more substantial disruption when compared to other populations (MSM facility testing was reduced by 59%, compared to 29% for the entire population [17]). Therefore, we may have underestimated the disruption to ART initiations. The survey data only gave semi-quantitative estimates of disruptions to partnerships and condom use i.e. proportion of MSM having fewer partners and not the overall reduction in partner numbers. We accounted for this by exploring uncertainty in the reduction and sampling from a wider distribution of estimates. Further, the disruption estimates for testing/ART initiations/VS came from Jiangsu, different to the cities we model (Guangdong/Shandong), and estimates for disruptions to partner numbers/condom use came from 31 provinces, meaning we may not have fully captured the impact of COVID-19 in each city. Finally, we have not modelled the direct impacts of COVID-19 infection. Future extensions work could include modelling the potential characteristics of co-infection between HIV and COVID-19.

## Conclusions

The COVID-19 emergency is impacting HIV care worldwide, as face-to-face consultations and laboratory testing are reduced, drug and condom manufacture and transport are interrupted, and lockdowns affect peoples’ ability to access testing or collect medicines. Gaps in HIV treatment could lead to increased deaths from HIV and further HIV transmission, placing further burdens upon healthcare systems. The overall impact of COVID-19 on new HIV infections and HIV-related deaths is expected to be low to moderate for MSM in China, but this is dependent on the scale and length of the various disruptions. Resources should be urgently directed to ensuring VS and condom use remain high in order to mitigate any adverse effects of COVID-19 disruption on HIV transmission and control among MSM in China.

## Data Availability

Data can be found in the supplement (Tables S1-S6)

## Authors’ contributions

Conception and design of the study: RDB, GF, JJO, JDT, KMET, WT, PV, KMM. Acquisition of data: GF, JL, WT. Mathematical modelling: RDB, LM, KMET, PV, KMM. Coding and simulations: RDB. Analysis and interpretation of results: RDB, GF, LM, JL, JJO, JDT, KMET, WT, PV, KMM. Writing and drafting of the manuscript: RDB, GF, LM, JL, JJO, JDT, KMET, WT, PV, KMM. Approval of the submitted manuscript: RDB, GF, LM, JL, JJO, JDT, KMET, WT, PV, KMM.

## Acknowledgements

### Funding

This work was supported by Global Public Health strand of the Elizabeth Blackwell Institute for Health Research, funded under the University of Bristol’s QR GCRF strategy (ISSF3: 204813/Z/16/Z). This work was also supported by the US NIH (NIAID K24AI143471, 1R01AI114310) and the NIHR Health Protection Research Unit in Behavioural Science and Evaluation at the University of Bristol (award number NIHR200877). This work was also supported by Health Data Research UK, which is funded by the UK Medical Research Council, Engineering and Physical Sciences Research Council, Economic and Social Research Council, National Institute for Health Research, Chief Scientist Office of the Scottish Government Health and Social Care Directorates, Health and Social Care Research and Development Division (Welsh Government), Public Health Agency (South Western Ireland), British Heart Foundation and Wellcome (award number CFC0129).

## Additional files

Additional file 1: Supplement **Supplement**.**docx**

Information on file format. Further details of the mathematical model and full results.

## List of abbreviations

HIV: human immunodeficiency virus
COVID-19: the disease caused by the SARS-CoV-2 (2019-nCoV) coronavirus.
PLWH: people living with HIV
MSM: men who have sex with men
ART: antiretroviral therapy
LMIC: low- and middle-income countries
VS: viral suppression
CrI: credible interval
CI: confidence interval
CDC: centre for disease control

