## supplement for "Estimating the impact of disruptions due to COVID-19 on HIV transmission and control among men who have sex with men in China": Supplement.docx

**Full results**

**Table S1 All cities combined**

| All cities change in new HIV infections | 1 year | | | | 5 year | | | |
| --- | --- | --- | --- | --- | --- | --- | --- | --- |
| Scenario (3M disruption unless otherwise stated) | Change in new HIV infections (95%CI) | | % Change in new HIV infections (95%CI) | | Change in new HIV infections (95%CI) | | % Change in new HIV infections (95%CI) | |
| A facility testing | 0.8 | (0.4, 2) | 2.3 | (1.7, 2.9) | 0.7 | (0.3, 1.5) | 1.7 | (1.2, 2.4) |
| B ART initiation | 0.7 | (0.3, 1.5) | 1.7 | (1.2, 2.4) | 2.0 | (0.9, 3.9) | 1.1 | (0.7, 1.8) |
| C partnerships | -6.2 | (-16, -2.4) | -16.2 | (-23.2, -11.1) | -9.5 | (-25.2, -3.5) | -5.3 | (-8.1, -3.5) |
| D condom use | 3.1 | (1.1, 8) | 7.8 | (4.5, 13.8) | 4.6 | (1.6, 11.5) | 2.6 | (1.4, 4.7) |
| A+B+C+D | -3.2 | (-11, -0.7) | -8.7 | (-17.2, -2.8) | -2.7 | (-12.7, 1.1) | -1.6 | (-4.3, 0.6) |
| E10 viral suppression | 1.2 | (0.6, 2.4) | 3.0 | (1.9, 5.6) | 1.8 | (0.8, 3.4) | 0.9 | (0.6, 1.9) |
| E25 viral suppression | 3.1 | (1.4, 5.9) | 7.4 | (4.7, 14) | 4.4 | (1.9, 8.5) | 2.4 | (1.4, 4.8) |
| A+B+C+D+E10 | -2.5 | (-10, 0) | -6.6 | (-15.9, 0) | -1.6 | (-11.3, 2.4) | -0.9 | (-3.8, 1.5) |
| A+B+C+D+E25 | -1.5 | (-8.9, 1.9) | -3.6 | (-13.9, 5) | 0.1 | (-9.3, 5.1) | 0.1 | (-3.4, 3.1) |
| A+B+C+D (4 months) | -4.2 | (-14.5, -0.9) | -11.7 | (-22.4, -3.8) | -3.5 | (-16.6, 1.5) | -2.2 | (-5.5, 0.8) |
| A+B+C+D (6 months) | -6.4 | (-21.5, -1.4) | -17.7 | (-32.4, -6.1) | -5.1 | (-24.4, 2.2) | -3.1 | (-7.9, 1.1) |
| A+B+C+D+E25 (4 months) | -2.0 | (-11.7, 2.4) | -4.9 | (-18.1, 6.5) | 0.2 | (-12.2, 6.8) | 0.1 | (-4.4, 4.1) |
| A+B+C+D+E25 (6 months) | -3.1 | (-17.3, 3.3) | -7.5 | (-26.1, 9) | 0.3 | (-17.9, 10.2) | 0.2 | (-6.4, 6.1) |

| All cities change in HIV-related deaths | 1 year | | | | 5 year | | | |
| --- | --- | --- | --- | --- | --- | --- | --- | --- |
| Scenario (3M disruption unless otherwise stated) | Change in HIV-related deaths (95%CI) | | % Change in HIV-related deaths (95%CI) | | Change in HIV-related deaths (95%CI) | | % Change in HIV-related deaths (95%CI) | |
| A facility testing | 0.0 | (0, 0.1) | 0.2 | (0.1, 0.3) | 0.4 | (0.2, 0.6) | 1.8 | (1.5, 2) |
| B ART initiation | 0.4 | (0.2, 0.6) | 1.8 | (1.5, 2) | 1.2 | (0.7, 1.9) | 1.3 | (1, 1.5) |
| C partnerships | -0.1 | (-0.3, -0.1) | -0.6 | (-1, -0.4) | -1.5 | (-2.8, -0.7) | -1.5 | (-2.3, -1) |
| D condom use | 0.1 | (0, 0.1) | 0.3 | (0.1, 0.5) | 0.7 | (0.3, 1.5) | 0.7 | (0.4, 1.2) |
| A+B+C+D | 0.3 | (0.2, 0.5) | 1.5 | (1.1, 1.8) | 0.6 | (-0.3, 1.4) | 0.6 | (-0.3, 1.2) |
| E10 viral suppression | 0.7 | (0.5, 1.1) | 3.5 | (2.6, 4.5) | 0.9 | (0.6, 1.3) | 0.9 | (0.7, 1.3) |
| E25 viral suppression | 1.8 | (1.2, 2.8) | 8.7 | (6.6, 11.2) | 2.3 | (1.5, 3.3) | 2.3 | (1.7, 3.3) |
| A+B+C+D+E10 | 1.0 | (0.7, 1.6) | 5.0 | (3.8, 6.2) | 1.4 | (0.4, 2.5) | 1.5 | (0.4, 2.2) |
| A+B+C+D+E25 | 2.1 | (1.4, 3.3) | 10.1 | (7.6, 12.7) | 2.6 | (1.5, 4) | 2.7 | (1.4, 3.9) |
| A+B+C+D (4 months) | 0.4 | (0.2, 0.7) | 2.0 | (1.5, 2.4) | 0.8 | (-0.4, 1.9) | 0.9 | (-0.3, 1.6) |
| A+B+C+D (6 months) | 0.6 | (0.3, 0.9) | 2.7 | (2.1, 3.3) | 1.2 | (-0.4, 2.8) | 1.3 | (-0.4, 2.4) |
| A+B+C+D+E25 (4 months) | 2.8 | (1.8, 4.3) | 13.3 | (10.1, 16.8) | 3.5 | (2, 5.4) | 3.6 | (2, 5.2) |
| A+B+C+D+E25 (6 months) | 4.1 | (2.7, 6.4) | 19.6 | (15, 24.8) | 5.3 | (3, 8.1) | 5.4 | (3, 7.7) |

**Table S2 Guangzhou**

| Guangzhou change in new HIV infections | 1 year | | | | 5 year | | | |
| --- | --- | --- | --- | --- | --- | --- | --- | --- |
| Scenario (3 month disruption unless otherwise stated) | Change in new HIV infections (95%CI) | | % Change in new HIV infections (95%CI) | | Change in new HIV infections (95%CI) | | % Change in new HIV infections (95%CI) | |
| A facility testing | 2.8 | (1, 5) | 2.2 | (1.7, 2.8) | 2.1 | (1.1, 4) | 1.7 | (1.1, 2.5) |
| B ART initiation | 2.1 | (1.1, 4) | 1.7 | (1.1, 2.5) | 4.9 | (2.5, 9.6) | 1.0 | (0.6, 1.6) |
| C partnerships | -19.9 | (-42.5, -7.3) | -15.6 | (-21.9, -10.5) | -24.4 | (-53.5, -9.5) | -4.6 | (-7.1, -3) |
| D condom use | 9.3 | (3.9, 24.3) | 7.7 | (4.1, 13.9) | 12.2 | (4.9, 32) | 2.3 | (1.2, 4.4) |
| A+B+C+D | -8.7 | (-27.1, -1.1) | -8.0 | (-14.8, -1) | -5.8 | (-24.4, 8) | -1.1 | (-3.3, 1.1) |
| E10 viral suppression | 4.5 | (2.2, 7.5) | 3.5 | (1.9, 7.2) | 5.6 | (2.7, 9.8) | 1.0 | (0.5, 2.3) |
| E25 viral suppression | 11.1 | (5.5, 18.5) | 8.6 | (4.7, 17.8) | 13.9 | (6.8, 24.1) | 2.4 | (1.2, 5.7) |
| A+B+C+D+E10 | -6.4 | (-24.2, 3) | -5.4 | (-13.2, 2.8) | -1.4 | (-21, 13.8) | -0.3 | (-2.8, 2) |
| A+B+C+D+E25 | -2.0 | (-19.9, 10.8) | -1.6 | (-11.4, 9.3) | 3.5 | (-15.8, 22.1) | 0.6 | (-2.2, 4.2) |
| A+B+C+D (4 months) | -11.7 | (-35.8, -1.6) | -10.7 | (-19.5, -1.5) | -7.6 | (-32.1, 10.6) | -1.4 | (-4.3, 1.5) |
| A+B+C+D (6 months) | -18.1 | (-53, -2.7) | -16.4 | (-28.8, -2.8) | -11.1 | (-47.3, 15.9) | -2.1 | (-6.3, 2.3) |
| A+B+C+D+E25 (4 months) | -2.8 | (-26.3, 13.9) | -2.3 | (-15.1, 12.1) | 4.7 | (-20.7, 29.5) | 0.7 | (-2.9, 5.6) |
| A+B+C+D+E25 (6 months) | -4.3 | (-39.1, 19.6) | -3.9 | (-22.5, 17.4) | 7.1 | (-30.3, 44.1) | 1.2 | (-4.2, 8.4) |

| Guangzhou change in HIV-related deaths | 1 year | | | | 5 year | | | |
| --- | --- | --- | --- | --- | --- | --- | --- | --- |
| Scenario (3M disruption unless otherwise stated) | Change in HIV-related deaths (95%CI) | | % Change in HIV-related deaths (95%CI) | | Change in HIV-related deaths (95%CI) | | % Change in HIV-related deaths (95%CI) | |
| A facility testing | 0.2 | (0.1, 0.3) | 0.2 | (0.1, 0.3) | 1.4 | (0.9, 2) | 1.7 | (1.5, 1.9) |
| B ART initiation | 1.4 | (0.9, 2) | 1.7 | (1.5, 1.9) | 3.9 | (2.3, 5.8) | 1.1 | (0.9, 1.3) |
| C partnerships | -0.3 | (-0.6, -0.2) | -0.4 | (-0.6, -0.3) | -3.3 | (-6.5, -1.5) | -1.0 | (-1.4, -0.6) |
| D condom use | 0.2 | (0.1, 0.4) | 0.2 | (0.1, 0.4) | 1.6 | (0.8, 4.1) | 0.5 | (0.2, 0.9) |
| A+B+C+D | 1.3 | (0.8, 1.9) | 1.6 | (1.3, 1.9) | 2.8 | (1.2, 5.2) | 0.8 | (0.3, 1.3) |
| E10 viral suppression | 3.3 | (2.4, 4.7) | 4.1 | (3, 5.6) | 3.7 | (2.7, 5.1) | 1.1 | (0.8, 1.5) |
| E25 viral suppression | 8.3 | (6, 11.7) | 10.3 | (7.4, 13.9) | 9.3 | (6.7, 12.8) | 2.8 | (2, 3.8) |
| A+B+C+D+E10 | 4.5 | (3.3, 6.4) | 5.8 | (4.4, 7.2) | 6.2 | (4, 9.4) | 1.9 | (1.2, 2.5) |
| A+B+C+D+E25 | 9.4 | (6.8, 13.3) | 11.8 | (8.6, 15.4) | 11.4 | (7.8, 16.3) | 3.4 | (2.3, 4.5) |
| A+B+C+D (4 months) | 1.7 | (1, 2.4) | 2.0 | (1.7, 2.4) | 3.7 | (1.6, 6.9) | 1.1 | (0.5, 1.7) |
| A+B+C+D (6 months) | 2.3 | (1.5, 3.4) | 2.8 | (2.4, 3.3) | 5.6 | (2.5, 10.4) | 1.7 | (0.7, 2.6) |
| A+B+C+D+E25 (4 months) | 12.4 | (9, 17.6) | 15.6 | (11.4, 20.5) | 15.1 | (10.4, 21.6) | 4.5 | (3.1, 6) |
| A+B+C+D+E25 (6 months) | 18.3 | (13.3, 26) | 23.1 | (16.9, 30.2) | 22.7 | (15.6, 32.3) | 6.7 | (4.7, 8.9) |

**Table S3 Shenzhen**

| Shenzhen change in new HIV infections | 1 year | | | | 5 year | | | |
| --- | --- | --- | --- | --- | --- | --- | --- | --- |
| Scenario (3M disruption unless otherwise stated) | Change in new HIV infections (95%CI) | | % Change in new HIV infections (95%CI) | | Change in new HIV infections (95%CI) | | % Change in new HIV infections (95%CI) | |
| A facility testing | 10.1 | (6.1, 14.6) | 2.2 | (1.7, 2.8) | 6.3 | (3.8, 9.8) | 1.4 | (0.9, 2.1) |
| B ART initiation | 6.3 | (3.8, 9.8) | 1.4 | (0.9, 2.1) | 16.4 | (9.3, 26.7) | 0.8 | (0.5, 1.4) |
| C partnerships | -68.2 | (-112.2, -39.2) | -15.3 | (-20.1, -10) | -81.9 | (-138, -45.8) | -3.9 | (-5.8, -2.6) |
| D condom use | 32.3 | (16.3, 60.8) | 6.8 | (4.1, 13.1) | 38.7 | (19.1, 74.4) | 1.8 | (1, 3.7) |
| A+B+C+D | -36.6 | (-75.2, -3.7) | -8.7 | (-14.6, -0.8) | -18.3 | (-59.8, 21.6) | -1.0 | (-2.8, 1.3) |
| E10 viral suppression | 13.4 | (7.2, 20.9) | 2.9 | (1.5, 5.3) | 16.1 | (7.8, 26.2) | 0.8 | (0.4, 1.5) |
| E25 viral suppression | 33.2 | (17.7, 51.7) | 7.3 | (3.6, 13) | 39.8 | (19.2, 64.8) | 1.9 | (0.9, 3.7) |
| A+B+C+D+E10 | -27.9 | (-68, 8) | -7.1 | (-13.4, 2.1) | -9.4 | (-48.4, 40) | -0.5 | (-2.3, 2.1) |
| A+B+C+D+E25 | -17.4 | (-57.9, 28) | -3.9 | (-11, 6.9) | 6.6 | (-38, 65) | 0.4 | (-1.7, 3.7) |
| A+B+C+D (4 months) | -48.9 | (-100.1, -5.7) | -11.7 | (-19.4, -1.3) | -23.8 | (-78.8, 28.8) | -1.3 | (-3.6, 1.7) |
| A+B+C+D (6 months) | -74.0 | (-151.7, -11) | -17.8 | (-28.8, -2.5) | -35.6 | (-116.4, 43.1) | -2.0 | (-5.3, 2.6) |
| A+B+C+D+E25 (4 months) | -23.6 | (-77.7, 36.3) | -5.3 | (-14.6, 9) | 9.1 | (-50.1, 86.5) | 0.5 | (-2.2, 4.9) |
| A+B+C+D+E25 (6 months) | -36.3 | (-118.4, 51.1) | -8.3 | (-21.9, 12.7) | 14.2 | (-73.8, 129.4) | 0.8 | (-3.2, 7.4) |

| Shenzhen change in HIV-related deaths | 1 year | | | | 5 year | | | |
| --- | --- | --- | --- | --- | --- | --- | --- | --- |
| Scenario (3M disruption unless otherwise stated) | Change in HIV-related deaths (95%CI) | | % Change in HIV-related deaths (95%CI) | | Change in HIV-related deaths (95%CI) | | % Change in HIV-related deaths (95%CI) | |
| A facility testing | 0.4 | (0.2, 0.6) | 0.2 | (0.1, 0.2) | 4.0 | (2.7, 5.5) | 1.5 | (1.2, 1.6) |
| B ART initiation | 4.0 | (2.7, 5.5) | 1.5 | (1.2, 1.6) | 12.2 | (7.9, 18.9) | 1.0 | (0.8, 1.2) |
| C partnerships | -0.9 | (-1.4, -0.5) | -0.3 | (-0.5, -0.2) | -10.2 | (-18.1, -6.1) | -0.9 | (-1.3, -0.6) |
| D condom use | 0.4 | (0.2, 0.8) | 0.1 | (0.1, 0.3) | 4.9 | (2.4, 10.3) | 0.4 | (0.2, 0.7) |
| A+B+C+D | 3.6 | (2.5, 5.1) | 1.3 | (1.1, 1.6) | 8.9 | (3.6, 16.1) | 0.7 | (0.3, 1.2) |
| E10 viral suppression | 9.4 | (7.6, 11.9) | 3.5 | (2.6, 4.6) | 10.5 | (8.1, 13.8) | 0.9 | (0.6, 1.1) |
| E25 viral suppression | 23.4 | (18.9, 29.7) | 8.7 | (6.5, 11.5) | 26.2 | (20.2, 34.4) | 2.2 | (1.5, 2.8) |
| A+B+C+D+E10 | 12.9 | (10, 16.5) | 4.8 | (3.7, 5.9) | 18.5 | (11.6, 27.7) | 1.5 | (1, 2.1) |
| A+B+C+D+E25 | 26.8 | (21.4, 34) | 9.9 | (7.5, 12.7) | 33.1 | (23.1, 46) | 2.7 | (1.9, 3.7) |
| A+B+C+D (4 months) | 4.6 | (3.2, 6.5) | 1.7 | (1.4, 2.1) | 11.9 | (5, 21.4) | 0.9 | (0.5, 1.6) |
| A+B+C+D (6 months) | 6.3 | (4.4, 9) | 2.3 | (2, 2.9) | 18.1 | (7.8, 31.9) | 1.4 | (0.7, 2.4) |
| A+B+C+D+E25 (4 months) | 35.4 | (28.3, 45.1) | 13.1 | (10, 16.8) | 44.0 | (30.8, 61.2) | 3.6 | (2.6, 4.9) |
| A+B+C+D+E25 (6 months) | 52.4 | (41.9, 66.7) | 19.4 | (14.8, 24.9) | 65.8 | (46.1, 91.1) | 5.4 | (3.8, 7.3) |

**Table S4 Jinan**

| Jinan change in new HIV infections | 1 year | | | | 5 year | | | |
| --- | --- | --- | --- | --- | --- | --- | --- | --- |
| Scenario (3M disruption unless otherwise stated) | Change in new HIV infections (95%CI) | | % Change in new HIV infections (95%CI) | | Change in new HIV infections (95%CI) | | % Change in new HIV infections (95%CI) | |
| A facility testing | 0.8 | (0.4, 1.4) | 2.0 | (1.6, 2.6) | 0.6 | (0.4, 1) | 1.6 | (1.2, 2.4) |
| B ART initiation | 0.6 | (0.4, 1) | 1.6 | (1.2, 2.4) | 1.8 | (1, 2.8) | 1.0 | (0.6, 1.6) |
| C partnerships | -5.9 | (-11.3, -3) | -15.6 | (-20.1, -10.7) | -7.9 | (-15, -3.9) | -4.8 | (-6.4, -3.1) |
| D condom use | 2.6 | (1, 5.8) | 6.4 | (3.5, 12.8) | 3.4 | (1.2, 7.6) | 1.9 | (1, 4.1) |
| A+B+C+D | -3.2 | (-8, -0.9) | -9.1 | (-15, -2.5) | -2.6 | (-7.8, 1) | -1.5 | (-3.4, 0.5) |
| E10 viral suppression | 1.2 | (0.7, 1.9) | 3.0 | (1.8, 5.8) | 1.6 | (0.8, 2.6) | 0.9 | (0.5, 1.9) |
| E25 viral suppression | 3.0 | (1.7, 4.7) | 7.5 | (4.4, 14.5) | 3.9 | (2.1, 6.4) | 2.1 | (1.2, 4.8) |
| A+B+C+D+E10 | -2.8 | (-7.3, 0.3) | -7.3 | (-13.5, 0.8) | -1.7 | (-6.6, 2.5) | -1.0 | (-3, 1.5) |
| A+B+C+D+E25 | -1.7 | (-6.2, 1.9) | -4.4 | (-11.7, 6.9) | -0.3 | (-5.5, 5) | -0.1 | (-2.3, 3.6) |
| A+B+C+D (4 months) | -4.3 | (-10.6, -1.2) | -12.1 | (-19.7, -3.5) | -3.5 | (-10.2, 1.3) | -2.0 | (-4.4, 0.7) |
| A+B+C+D (6 months) | -6.3 | (-15.7, -2) | -18.3 | (-29, -5.7) | -5.1 | (-14.9, 2) | -3.0 | (-6.4, 1) |
| A+B+C+D+E25 (4 months) | -2.2 | (-8.2, 2.5) | -6.0 | (-15.4, 9) | -0.3 | (-7.3, 6.6) | -0.1 | (-3.1, 4.7) |
| A+B+C+D+E25 (6 months) | -3.4 | (-12.1, 3.5) | -9.3 | (-22.9, 12.7) | -0.4 | (-10.7, 9.9) | -0.2 | (-4.5, 7.1) |

| Jinan change in HIV-related deaths | 1 year | | | | 5 year | | | |
| --- | --- | --- | --- | --- | --- | --- | --- | --- |
| Scenario (3M disruption unless otherwise stated) | Change in HIV-related deaths (95%CI) | | % Change in HIV-related deaths (95%CI) | | Change in HIV-related deaths (95%CI) | | % Change in HIV-related deaths (95%CI) | |
| A facility testing | 0.0 | (0, 0.1) | 0.1 | (0.1, 0.2) | 0.4 | (0.3, 0.6) | 1.7 | (1.6, 1.9) |
| B ART initiation | 0.4 | (0.3, 0.6) | 1.7 | (1.6, 1.9) | 1.3 | (0.9, 2.1) | 1.2 | (1, 1.3) |
| C partnerships | -0.1 | (-0.2, -0.1) | -0.4 | (-0.6, -0.2) | -1.2 | (-2.2, -0.6) | -1.1 | (-1.5, -0.6) |
| D condom use | 0.0 | (0, 0.1) | 0.2 | (0.1, 0.4) | 0.5 | (0.2, 1) | 0.4 | (0.2, 0.9) |
| A+B+C+D | 0.4 | (0.3, 0.5) | 1.5 | (1.3, 1.7) | 0.8 | (0.2, 1.5) | 0.7 | (0.2, 1.2) |
| E10 viral suppression | 0.9 | (0.7, 1.2) | 3.6 | (3.2, 4.5) | 1.1 | (0.8, 1.4) | 1.0 | (0.8, 1.3) |
| E25 viral suppression | 2.3 | (1.7, 3.1) | 9.1 | (7.9, 11.1) | 2.7 | (2, 3.4) | 2.4 | (2, 3.2) |
| A+B+C+D+E10 | 1.3 | (0.9, 1.8) | 5.1 | (4.4, 6) | 1.8 | (1, 2.7) | 1.5 | (1, 2.2) |
| A+B+C+D+E25 | 2.7 | (2, 3.5) | 10.5 | (9.1, 12.5) | 3.3 | (2.1, 4.5) | 2.9 | (2.1, 3.9) |
| A+B+C+D (4 months) | 0.5 | (0.3, 0.7) | 1.9 | (1.7, 2.2) | 1.0 | (0.3, 2) | 0.9 | (0.3, 1.5) |
| A+B+C+D (6 months) | 0.7 | (0.5, 1) | 2.7 | (2.3, 3.1) | 1.6 | (0.5, 2.9) | 1.4 | (0.5, 2.3) |
| A+B+C+D+E25 (4 months) | 3.5 | (2.6, 4.7) | 13.9 | (12, 16.6) | 4.4 | (2.8, 6) | 3.8 | (2.8, 5.2) |
| A+B+C+D+E25 (6 months) | 5.2 | (3.9, 6.9) | 20.5 | (17.8, 24.5) | 6.5 | (4.3, 9) | 5.8 | (4.2, 7.8) |

**Table S5 Qingdao**

| Qingdao change in new HIV infections | 1 year | | | | 5 year | | | |
| --- | --- | --- | --- | --- | --- | --- | --- | --- |
| Scenario (3M disruption unless otherwise stated) | Change in new HIV infections (95%CI) | | % Change in new HIV infections (95%CI) | | Change in new HIV infections (95%CI) | | % Change in new HIV infections (95%CI) | |
| A facility testing | 0.8 | (0.4, 2) | 2.3 | (1.7, 2.9) | 0.7 | (0.3, 1.5) | 1.7 | (1.2, 2.4) |
| B ART initiation | 0.7 | (0.3, 1.5) | 1.7 | (1.2, 2.4) | 2.0 | (0.9, 3.9) | 1.1 | (0.7, 1.8) |
| C partnerships | -6.2 | (-16, -2.4) | -16.2 | (-23.2, -11.1) | -9.5 | (-25.2, -3.5) | -5.3 | (-8.1, -3.5) |
| D condom use | 3.1 | (1.1, 8) | 7.8 | (4.5, 13.8) | 4.6 | (1.6, 11.5) | 2.6 | (1.4, 4.7) |
| A+B+C+D | -3.2 | (-11, -0.7) | -8.7 | (-17.2, -2.8) | -2.7 | (-12.7, 1.1) | -1.6 | (-4.3, 0.6) |
| E10 viral suppression | 1.2 | (0.6, 2.4) | 3.0 | (1.9, 5.6) | 1.8 | (0.8, 3.4) | 0.9 | (0.6, 1.9) |
| E25 viral suppression | 3.1 | (1.4, 5.9) | 7.4 | (4.7, 14) | 4.4 | (1.9, 8.5) | 2.4 | (1.4, 4.8) |
| A+B+C+D+E10 | -2.5 | (-10, 0) | -6.6 | (-15.9, 0) | -1.6 | (-11.3, 2.4) | -0.9 | (-3.8, 1.5) |
| A+B+C+D+E25 | -1.5 | (-8.9, 1.9) | -3.6 | (-13.9, 5) | 0.1 | (-9.3, 5.1) | 0.1 | (-3.4, 3.1) |
| A+B+C+D (4 months) | -4.2 | (-14.5, -0.9) | -11.7 | (-22.4, -3.8) | -3.5 | (-16.6, 1.5) | -2.2 | (-5.5, 0.8) |
| A+B+C+D (6 months) | -6.4 | (-21.5, -1.4) | -17.7 | (-32.4, -6.1) | -5.1 | (-24.4, 2.2) | -3.1 | (-7.9, 1.1) |
| A+B+C+D+E25 (4 months) | -2.0 | (-11.7, 2.4) | -4.9 | (-18.1, 6.5) | 0.2 | (-12.2, 6.8) | 0.1 | (-4.4, 4.1) |
| A+B+C+D+E25 (6 months) | -3.1 | (-17.3, 3.3) | -7.5 | (-26.1, 9) | 0.3 | (-17.9, 10.2) | 0.2 | (-6.4, 6.1) |

| Qingdao change in HIV-related deaths | 1 year | | | | 5 year | | | |
| --- | --- | --- | --- | --- | --- | --- | --- | --- |
| Scenario (3M disruption unless otherwise stated) | Change in HIV-related deaths (95%CI) | | % Change in HIV-related deaths (95%CI) | | Change in HIV-related deaths (95%CI) | | % Change in HIV-related deaths (95%CI) | |
| A facility testing | 0.0 | (0, 0.1) | 0.2 | (0.1, 0.3) | 0.4 | (0.2, 0.6) | 1.8 | (1.5, 2) |
| B ART initiation | 0.4 | (0.2, 0.6) | 1.8 | (1.5, 2) | 1.2 | (0.7, 1.9) | 1.3 | (1, 1.5) |
| C partnerships | -0.1 | (-0.3, -0.1) | -0.6 | (-1, -0.4) | -1.5 | (-2.8, -0.7) | -1.5 | (-2.3, -1) |
| D condom use | 0.1 | (0, 0.1) | 0.3 | (0.1, 0.5) | 0.7 | (0.3, 1.5) | 0.7 | (0.4, 1.2) |
| A+B+C+D | 0.3 | (0.2, 0.5) | 1.5 | (1.1, 1.8) | 0.6 | (-0.3, 1.4) | 0.6 | (-0.3, 1.2) |
| E10 viral suppression | 0.7 | (0.5, 1.1) | 3.5 | (2.6, 4.5) | 0.9 | (0.6, 1.3) | 0.9 | (0.7, 1.3) |
| E25 viral suppression | 1.8 | (1.2, 2.8) | 8.7 | (6.6, 11.2) | 2.3 | (1.5, 3.3) | 2.3 | (1.7, 3.3) |
| A+B+C+D+E10 | 1.0 | (0.7, 1.6) | 5.0 | (3.8, 6.2) | 1.4 | (0.4, 2.5) | 1.5 | (0.4, 2.2) |
| A+B+C+D+E25 | 2.1 | (1.4, 3.3) | 10.1 | (7.6, 12.7) | 2.6 | (1.5, 4) | 2.7 | (1.4, 3.9) |
| A+B+C+D (4 months) | 0.4 | (0.2, 0.7) | 2.0 | (1.5, 2.4) | 0.8 | (-0.4, 1.9) | 0.9 | (-0.3, 1.6) |
| A+B+C+D (6 months) | 0.6 | (0.3, 0.9) | 2.7 | (2.1, 3.3) | 1.2 | (-0.4, 2.8) | 1.3 | (-0.4, 2.4) |
| A+B+C+D+E25 (4 months) | 2.8 | (1.8, 4.3) | 13.3 | (10.1, 16.8) | 3.5 | (2, 5.4) | 3.6 | (2, 5.2) |
| A+B+C+D+E25 (6 months) | 4.1 | (2.7, 6.4) | 19.6 | (15, 24.8) | 5.3 | (3, 8.1) | 5.4 | (3, 7.7) |

**Mathematical model (from supplement of Booton et al** ^1^**)**

**Table S6**

| **Parameters** | **Guangzhou** | **Shenzhen** | **Jinan** | **Qingdao** | **Source** |
| --- | --- | --- | --- | --- | --- |
| *Sexual behaviour parameters* | | | | | |
| Average number of partners per year | | | | |  |
| Low risk, always insertive | 1.6 to 2.6 | 1.6 to 2.6 | 1.3 to 2.2 | 1.6 to 2.7 | Baseline trial data ^2^ |
| Low risk, versatile | 1.4 to 2.1 | 1.6 to 2.3 | 1.4 to 2.2 | 1.5 to 2.2 | Baseline trial data ^2^ |
| Low risk, always receptive | 1.2 to 2.2 | 1.2 to 2.3 | 1.4 to 2.5 | 1.7 to 2.8 | Baseline trial data ^2^ |
| High risk, always insertive | 11.8 to 18.9 | 14.6 to 22.4 | 3.6 to 22.4 | 12.3 to 24.0 | Baseline trial data ^2^ |
| High risk, versatile | 12.4 to 19.9 | 12.5 to 20.8 | 9.3 to 19.5 | 12.6 to 22.9 | Baseline trial data ^2^ |
| High risk, always receptive | 11.2 to 19.9 | 10.0 to 27.2 | 7.6 to 20.7 | 10.7 to 23.9 | Baseline trial data ^2^ |
| Anal sex acts per MSM partnership per year (assumed same for all cities) | | | | | |
| Low risk | 17 to 22 | | | | Baseline trial data ^2^ |
| High risk | 11 to 16 | | | | Baseline trial data ^2^ |
| Percentage of sex acts in which a condom is used | | |  |  |  |
| Low risk | 64 to 81 | 68 to 83 | 64 to 80 | 67 to 82 | Baseline trial data ^2^ |
| High risk | 73 to 92 | 67 to 89 | 61 to 88 | 60 to 87 | Baseline trial data ^2^ |
| *Intervention parameters* |  |  |  |  |  |
| Initial rate of facility-based testing per year | 0.1 to 0.2 (2005) | 0.4 to 0.6 (2006) | 0.3 to 0.4 (2005.5) | 0.3 to 0.4 (2006) | City level estimates ^3–7^ |
| Annual testing rates |  |  |  |  |  |
| First self-test, low risk | 0.14 to 0.30 | 0.07 to 0.15 | 0.19 to 0.23 | 0.10 to 0.30 | Baseline trial data ^2^ |
| First facility test, low risk | 0.10 to 0.20 | | | | Baseline trial data ^2^ |
| Self-test if not tested last 3 months | 0.1 to 0.2 | 0.1 to 0.2 | 0.1 to 0.3 | 0.2 to 0.4 | Baseline trial data ^2^ |
| Self-test, if tested in last 3 months | 0.7 to 1.4 | 0.7 to 1.2 | 0.7 to 1.4 | 0.6 to 0.9 | Baseline trial data ^2^ |
| Overall facility testing | 0.5 to 0.6 | | | | Baseline trial data ^2^ |
| Relative risk ratio for testing among high-risk MSM versus low-risk MSM | | | | | |
| Self-testing | 1.1 to 1.5 | | | | Baseline trial data ^2^ |
| Facility testing | 1.2 to 1.5 | | | | Baseline trial data ^2^ |
| Rate of dropout from ART per year | 0.02 to 0.04 | | | | National and Guangdong ^8,9^ |
| **Fitting metrics** | **Guangzhou** | **Shenzhen** | **Jinan** | **Qingdao** | **Source** |
| Size of MSM population at two time points | 12249 to 49698  (2008)  38570 to 57856  (2011)* | 45801 to 107082  (2006)*  120000 to 180000 (2016)* | 9687 to 19373 (2009)  12676 to 18830 (2017) | 29300 to 46200 (2016)  28000 to 51000 (2018) | ^10,11^**,** Shenzhen CDC 2016, ^12^, Shandong CDC 2019 |
| HIV prevalence (%), 95% CI |  |  |  |  |  |
| 2005 | N.D. | 0.0 to 3.3 | N.D. | N.D. | SZ CDC |
| 2006 | 0.4 to 2.9 | 1.0 to 3.8 | N.D. | N.D. | GZ, SZ CDC, ^13^ |
| 2007 | N.D. | 2.9 to 5.9 | N.D. | N.D. | SZ CDC |
| 2008 | 2.3 to 7.2 | 5.2 to 8.5 | N.D. | N.D. | GZ, SZ CDC ^14^ |
| 2009 | 2.0 to 5.8 | 7.2 to 10.9 | N.D. | N.D. | GZ, SZ CDC ^14^ |
| 2010 | 5.0 to 10.2 | 6.1 to 9.0 | 1.2 to 2.9 | 0.0 to 1.2 | GZ, SZ, SD CDC ^14^ |
| 2011 | 6.4 to 12.1 | 5.3 to 8.9 | 3.9 to 6.2 | 0.0 to 1.2 | GZ, SZ, SD CDC ^14,15^ |
| 2012 | 7.0 to 12.9 | 7.2 to 12.4 | 5.9 to 7.8 | 0.0 to 1.6 | GZ, SZ, SD CDC ^14,15^ |
| 2013 | 8.9 to 13.8 | 7.6 to 12.1 | 9.5 to 12.1 | 2.3 to 6.5 | GZ, SZ, SD CDC ^14,15^ |
| 2014 | 9.7 to 14.9 | 10.9 to 16.4 | 10.9 to 13.5 | 3.3 to 5.6 | GZ, SZ, SD CDC ^15,16^ |
| 2015 | 8.5 to 13.5 | 6.5 to 11.6 | 7.1 to 9.5 | 2.2 to 3.8 | GZ, SZ, SD CDC ^15,16^ |
| 2016 | N.D. | 9.1 to 16.0 | N.D. | N.D. | SZ CDC, ^15^ |
| HIV incidence rate (per 100 person-years) | 2.9 to 8.7 (2009.5) | 4.6 to 11.9 (2010) | N.D. | N.D. | ^17,18^ |
| Percentage of infected MSM diagnosed, 2013 | 38 to 70 | | | | Shandong province estimate ^19^ |
| Percentage of diagnosed MSM on ART | | | | |  |
| 2005 | 18.9 to 40.8 | | | | National estimate ^20^ |
| 2007 | 20.5 to 37.1 | | | | National estimate ^20^ |
| 2009 | 26.7 to 40.0 | | | | National estimate ^20^ |
| 2011 | 38.0 to 49.6 | | | | National estimate ^20^ |
| 2013 | 50.7 to 60.4 | | | | National estimate ^20^ |
| 2015 | 62.9 to 70.6 | | | | National estimate ^20^ |

GZ = Guangzhou, SZ = Shenzhen, JN = Jinan, QD = Qingdao,

SD = Shandong, CDC = Center for Disease Control, N.D. = no data,

* indicates ± 20% on lower and upper bounds

**Table S6:** Key parameters and fitting metrics used in the model in Booton et al.^1^ (summary of 95% confidence interval uncertainty ranges) for Guangzhou, Shenzhen, Jinan and Qingdao cities.

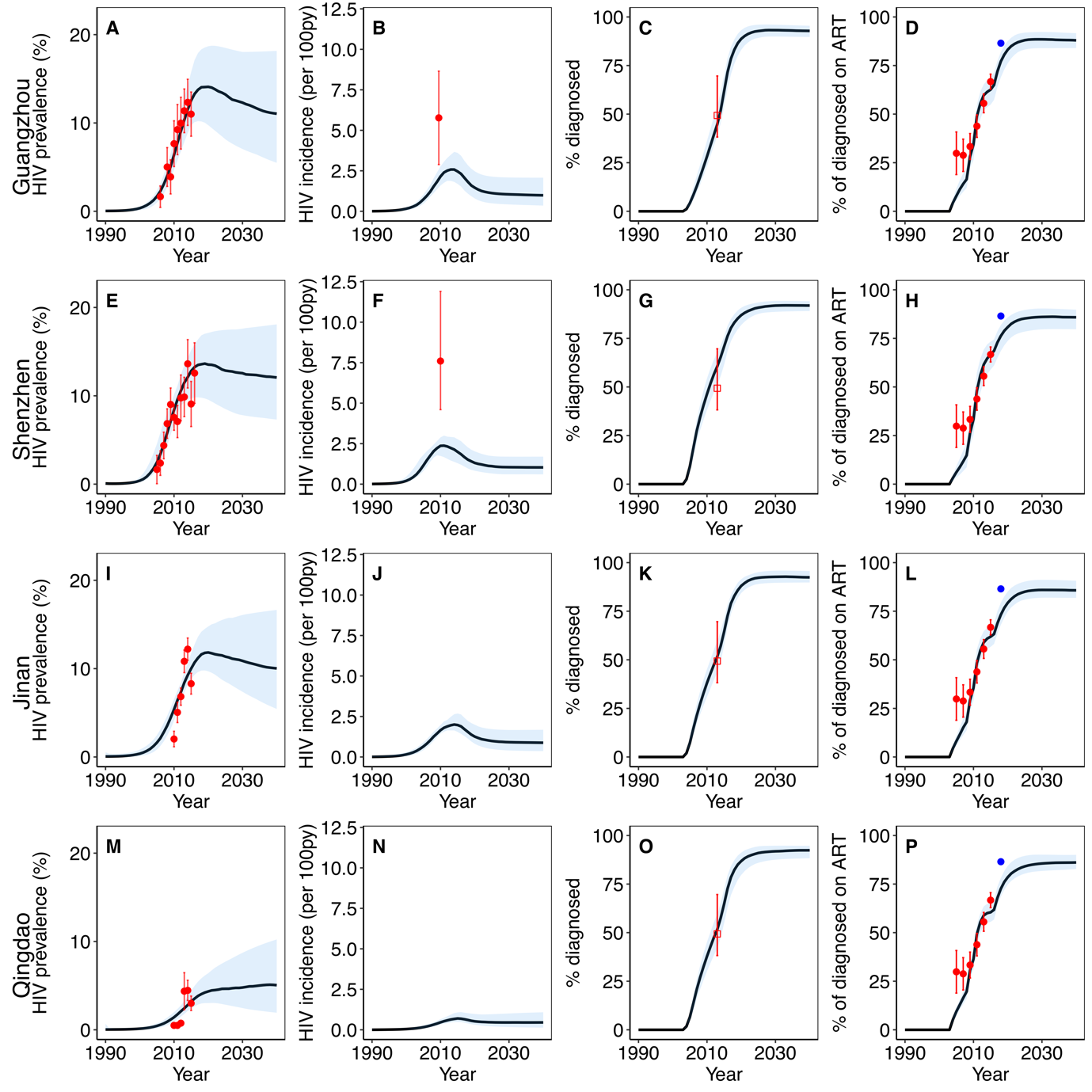
**Figure S1:** Comparison of the model projections in Booton et al.^1^ for each city against data on HIV prevalence, incidence per 100 person years, percentage on ART if diagnosed and percentage diagnosed. Projections are shown for Guangzhou (A-D), Shenzhen (E-H), Jinan (I-L) and Qingdao (M-P), with median (black line) and 95% credible interval (blue shaded area) being displayed for 100 model fits for each city. Empty red squares indicate HIV diagnosis data which was fit to (accepted if confidence interval) and red circles indicate those data which are included in the likelihood estimation to determine the best fitting model runs. The blue dot represents validation data from 2018 (86.5% of diagnosed MSM on ART in a recently published UNAIDS report ^21^)

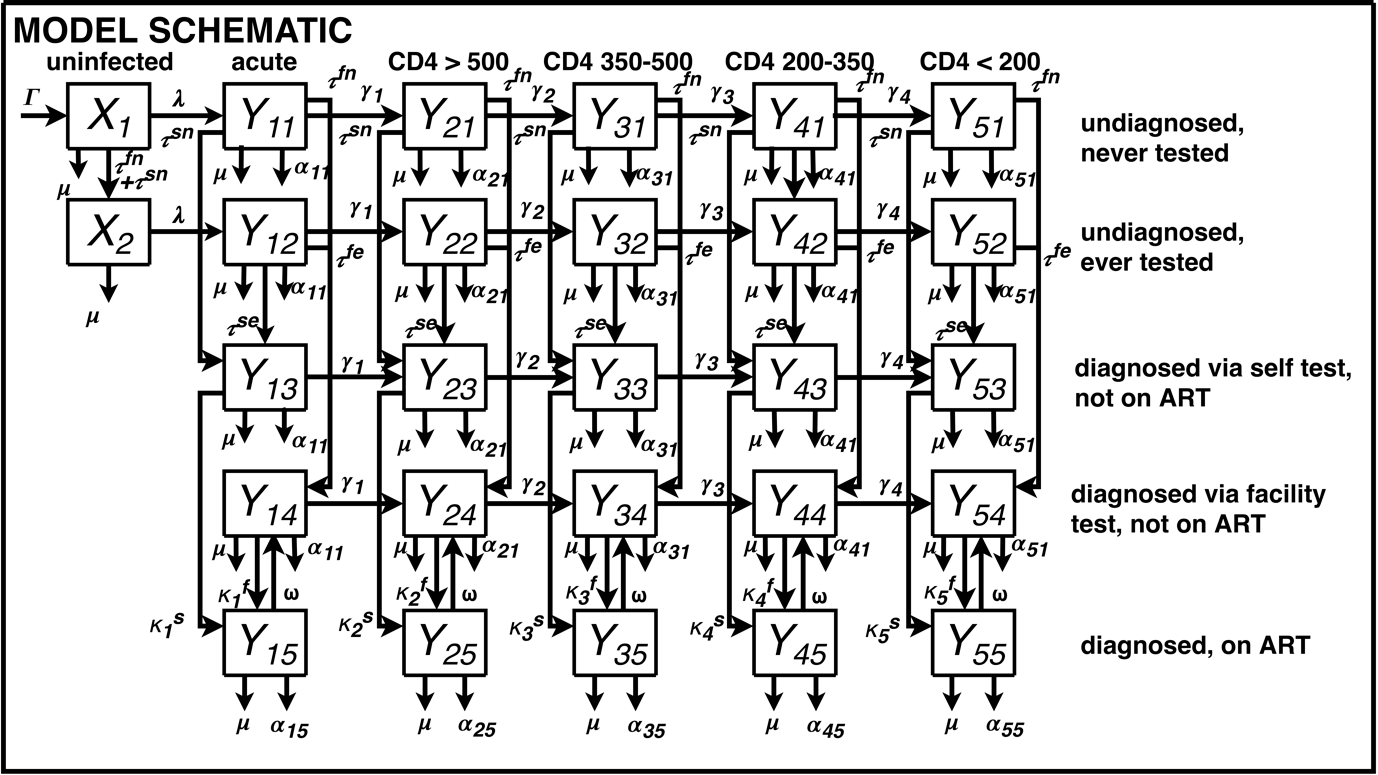

**Figure S2**: Mathematical model schematic for Booton et al.^1^. Subscripts on uninfected (X) stages indicate diagnosis/ART stage (y); subscripts on infected (Y) stages indicate infection stage (x) and diagnosis/ART stage (y). Risk and role subscripts are omitted for clarity.

Uninfected MSM are denoted by $X_{vwy}$, and infected MSM by $Y_{vwxy}$. Subscripts refer to $v=$risk (1= two or fewer male anal sex partners in the last 3 months, 2= three or more male anal sex partners in the last 3 months), $w=$role behaviour (1= always insertive, 2= versatile, 3= always receptive), $x$ is stage of infection (1= acute, 2= CD4>500, 3= CD4 351-500, 4= CD4 200-350, 5= CD4<200 cells/µl) and $y$ is diagnosis/ART (1= undiagnosed, never tested , 2= undiagnosed, ever tested, 3= diagnosed via self-test, not on ART, 4= diagnosed via facility test, not on ART and 5= diagnosed, on ART).

New MSM join the sexually active MSM population on sexual debut at a rate $\Gamma$, which is further stratified according to the percentage of individuals within each risk/role group ($m_{vw}$) and individuals do not move between role and risk groups. The population of MSM in each city grows in line with reported estimates of MSM population size taken from provincial Chinese Centers for Disease Control (CDC) datasets (Table 1). The mathematical model schematic is shown in Figure S1.

Susceptible uninfected MSM become infected with HIV at a rate $\lambda_{v,w}$ and move into one of the acute undiagnosed stages $Y_{vw11}$ and $Y_{vw12}$, dependent on their never/ever testing status. Without treatment, MSM move into progressively lower CD4 categories over time. The rate of progression $\gamma_{x}$ determines the speed at which individuals move from disease stage $Y_{vwxy}$to $Y_{vw x+1 y}$. We do not explicitly model changes in CD4 for those on ART. We divide the death rate from each compartment into the non-HIV related death/ leaving rate $\mu$ and the HIV-related death rate from each compartment$\alpha_{xy}$. The non-HIV related death rate accounts for MSM leaving the active sexual population and applies equally to all compartments regardless of infection or diagnosis status. The HIV-related death rate applies to those compartments which contain infected MSM, and differs according to ART status and infection stage, with MSM with CD4<200 cells/µl having a much larger risk of mortality (HIV-related mortality is a function of CD4 count at ART initiation).

Uninfected individuals who test for the first time move from the uninfected never tested to the uninfected ever tested compartment. Infected individuals who have never previously tested undergo first HIV facility-based testing at rate $\tau_{v}^{fn}$ and first HIV self-testing at rate $\tau_{v}^{sn}$ and move into respective diagnosed via facility test, or diagnosed by self-test compartments, where MSM who are initially diagnosed are not immediately on ART. Likewise, $\tau_{v}^{fe}$ and $\tau_{v}^{se}$ represent the repeat testing rates for MSM who have ever tested via facility or self-test respectively. These four testing rates are stratified by risk (we identified differences between risk groups for all testing rates) and further the self-testing is city specific. $\kappa_{x}^{s}$ and $\kappa_{x}^{f}$ determine the rate of treatment initiation for those who are diagnosed via self-test or facility-based test for each CD4 count, moving into compartments for MSM who have been diagnosed, and are on ART. Those on ART have reduced mortality, which is a function of their CD4 count at ART initiation. $\omega$ determines the rate of drop out from ART and these MSM move back into the diagnosed via facility test, not on ART compartment.

HIV transmission via anal sex occurs between MSM at a rate dependent on the probability per receptive anal sex act of acquiring HIV from a HIV positive partner ^22,23^, role-specific relative risks of acquiring HIV ^22–24^, HIV prevalence, disease stage and ART coverage among partners, levels of viral suppression among those on ART, the number of partners per year ^2^, the average number of anal sex acts per partnership ^2^, the efficacy of condom use per-sex act ^25^, the proportion of sex acts in which a condom is used ^2^, and the relative infectiousness of an individual in each disease stage ^26–28^. The number of anal sex acts per MSM partnership per year is assumed to be the same across all three role groups, as there was no statistical difference between the trial data estimates for role. Those MSM living with HIV who are virally suppressed are presumed to be untransmittable (U=U) ^29^.

*Model equations (ODEs)*

*Undiagnosed, never tested* ($y=1$)

uninfected

$$\frac{dX_{vw1}}{dt}= \Gamma m_{vw}-\lambda_{vw}X_{vw1}-X_{vw1}(\mu+\tau_{v}^{fn}+\tau_{v}^{sn})$$

acute $x=1$

$$\frac{dY_{vw11}}{dt}= \lambda_{vw}X_{vw1}- Y_{vw11}(\mu+\alpha_{11}+\gamma_{1}+\tau_{v}^{fn}+\tau_{v}^{sn})$$

CD4 200-500 $y=2,3,4$

$$\frac{dY_{vwx1}}{dt}= \gamma_{x-1}Y_{vw x-1 1}- Y_{vwx1}(\mu+\alpha_{x1}+\gamma_{x}+\tau_{v}^{fn}+\tau_{v}^{sn})$$

CD4 <200 $x=5$

$$\frac{dY_{vw51}}{dt}= \gamma_{4}Y_{vw41}- Y_{vw51}(\mu+\alpha_{51}+ \tau_{v}^{fn}+\tau_{v}^{sn})$$

*Undiagnosed, ever tested* ($y=2$)

uninfected

$$\frac{dX_{vw2}}{dt}=(\tau_{v}^{fn}+\tau_{v}^{sn})X_{vw1}-\lambda_{vw}X_{vw2}-{\mu X}_{vw2}$$

acute $x=1$

$$\frac{dY_{vw12}}{dt}= \lambda_{vw}X_{vw2}- Y_{vw12}(\mu+\alpha_{11}+\gamma_{1}+\tau_{v}^{fe}+\tau_{v}^{se})$$

CD4 200-500 $y=2,3,4$

$$\frac{dY_{vwx2}}{dt}= \gamma_{x-1}Y_{vw x-1 2}- Y_{vwx2}(\mu+\alpha_{x1}+\gamma_{x}+\tau_{v}^{fe}+\tau_{v}^{se})$$

CD4 <200 $x=5$

$$\frac{dY_{vw52}}{dt}= \gamma_{4}Y_{vw42}- Y_{vw52}(\mu+\alpha_{51}+ \tau_{v}^{fe}+\tau_{v}^{se})$$

*Diagnosed via self-test, not on ART* ($y=3$)

acute $x=1$

$$\frac{dY_{vw13}}{dt}=\tau_{v}^{sn}Y_{vw11}+\tau_{v}^{se}Y_{vw12}- Y_{vw12}(\mu+\alpha_{11}+\gamma_{1}+\kappa_{1}^{s})$$

CD4 200-500 $x=2,3,4$

$$\frac{dY_{vwx3}}{dt}= \gamma_{x-1}Y_{vw x-1 3}+\tau_{v}^{sn}Y_{vwx1}+\tau_{v}^{se}Y_{vwx2}- Y_{vwx3}(\mu+\alpha_{x1}+\gamma_{x}+\kappa_{x}^{s})$$

CD4 <200 $x=5$

$$\frac{dY_{vw53}}{dt}= \gamma_{4}Y_{vw43}+\tau_{v}^{sn}Y_{vw51}+\tau_{v}^{se}Y_{vw52}- Y_{vw53}(\mu+\alpha_{51}+\kappa_{5}^{s})$$

*Diagnosed via facility-based test, not on ART* ($y=4$)

acute $x=1$

$$\frac{dY_{vw14}}{dt}= \tau_{v}^{fn}Y_{vw11}+\tau_{v}^{fe}Y_{vw12}+\omega Y_{vw15}- Y_{vw14}(\mu+\alpha_{11}+\gamma_{1}+\kappa_{1}^{f})$$

CD4 200-500 $x=2,3,4$

$$\frac{dY_{vwx4}}{dt}= \gamma_{x-1}Y_{vw x-1 4}+\tau_{v}^{fn}Y_{vwx1}+\tau_{v}^{fe}Y_{vwx2}+\omega Y_{vwx4}- Y_{vwx4}(\mu+\alpha_{x1}+\gamma_{x}+\kappa_{x}^{f})$$

CD4 <200 $x=5$

$$\frac{dY_{vw54}}{dt}= \gamma_{4}Y_{vw44}+\tau_{v}^{fn}Y_{vw51}+\tau_{v}^{fe}Y_{vw52}+\omega Y_{vw55}- Y_{vw54}(\mu+\alpha_{51}+\kappa_{5}^{f})$$

*Diagnosed, on ART* ($y=5$)

acute $x=1$ & CD4 200-500 $x=2,3,4$ & CD4<200 $x=5$

$$\frac{dY_{vwx5}}{dt}= \kappa_{x}^{s}Y_{vwx3}+\kappa_{x}^{f}Y_{vwx4}- Y_{vwx5}(\mu+\alpha_{x5}+\omega)$$

*Force of infection*

$\lambda_{vw}$ is the rate at which susceptible individuals in risk group $v$ and role group $w$ become infected and move into the acute $Y_{vw1y}$ compartment. This is the sum of each infection from group $Y_{v'w'xy}$ to $X_{vwy}$. $e_{c}$ is the efficacy of condom use per-sex-act, $s_{vwv'w'}$ is the proportion of sex acts in which a condom is used between groups $vw$ and $v'w'$. $\beta$ represents the probability per receptive-sex act of acquiring HIV from a HIV-positive partner, and $h_{w}$ is the relative risk of acquiring HIV when in role group $w$, compared to the risk when receptive. The relative risk for versatile MSM depends on their partner, e.g. if receptive, then versatile MSM will be the insertive partner. If versatile MSM partner with other versatile MSM we assume an equal number of insertive and receptive sex acts. $\rho_{vwv'w'}$ represents the proportion of partners who are in risk/role group $v'w'$ for those in group $vw$. $c_{vw}$ represents the number of partners per year for those in risk/role group $vw$. $n_{vw}$ is the average number of anal sex acts per partnership per year, which varies by risk group. $d_{xy}$ is the relative infectiousness of each disease stage and ART status (where $d_{x1}, d_{x2}, d_{x3}, d_{x4}$ is off ART and $d_{x5}$ is on ART) compared to an individual in the chronic CD4>200 off ART stage. We calculate the force of infection as

$$\lambda_{vw}= 1-\prod_{v^{'}=1}^{2} \prod_{w^{'}=1}^{3} \left( \frac{X_{v^{'}w^{'}}}{N_{v^{'}w^{'}}}+\sum_{y=1}^{5} \sum_{x=1}^{5} \frac{Y_{v^{'}w^{'}xy}}{N_{v^{'}w^{'}}}\left( 1-h_{w}d_{xy}\beta\left( 1-e_{c}s_{{vwv}^{'}w^{'}} \right) \right)^{n_{vw}} \right)^{\rho_{{vwv}^{'}w^{'}}c_{vw}}$$

Where the total number of MSM partners in group $v'w'$ is calculated as

$$N_{v^{'}w^{'}}= X_{v^{'}w^{'}}+\sum_{y=1}^{5} \sum_{x=1}^{5} Y_{v'w'xy}$$

and the probability $\rho_{vwv'w'}$ that an individual in risk group $v$ will choose a partner in risk group $v'$ as $\rho_{vv^{'}}$ using the following mixing matrix varying between assortative and proportionate mixing, and independent mixing assumed by risk and fixed role mixing, $\epsilon_{v}$ representing the probability of assortative mixing by risk: Mixing between MSM is assumed to be fixed by role (insertive, versatile and receptive) and is assumed to vary between assortative (like-with-like mixing; low with low, high with high risk) and proportionate (random mixing) by risk.

| Overall mixing varying by $\epsilon_{v}$ | Low risk | High risk |
| --- | --- | --- |
| Low risk | $\epsilon_{v}+\left( 1-\epsilon_{v} \right)Total_{low}$ | $\left( 1-\epsilon_{v} \right)Total_{high}$ |
| High risk | $\left( 1-\epsilon_{v} \right)Total_{low}$ | $\epsilon_{v}+\left( 1-\epsilon_{v} \right)Total_{high}$ |

An illustration of the sexual mixing used in this model is in Figure S2. Setting $\epsilon_{v}=0$represents fully proportionate mixing, and $\epsilon_{v}=1$ represents fully assortative mixing.

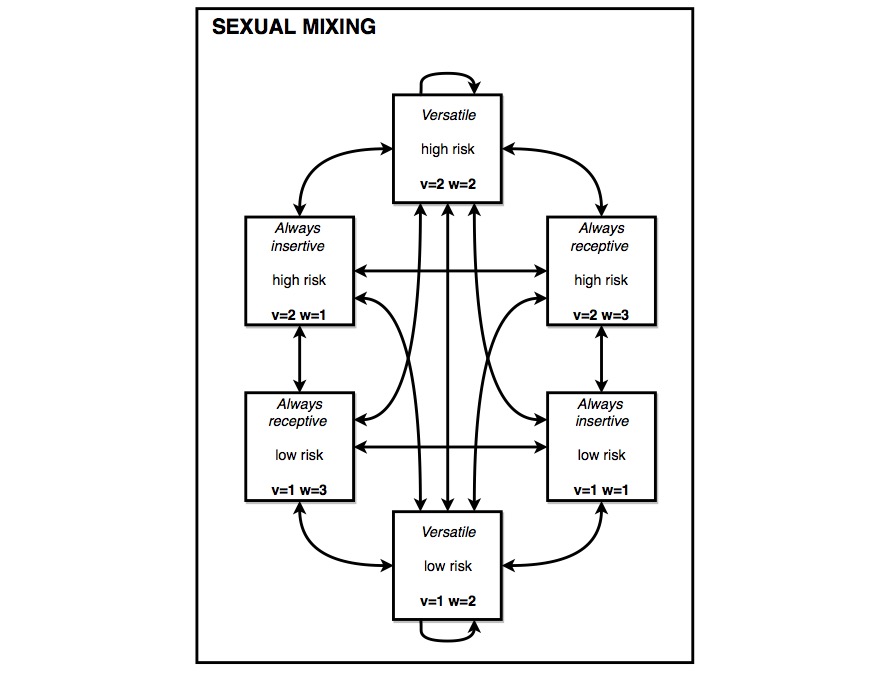

Figure S3: Sexual mixing with six groups by role and risk in Booton et al.^1^

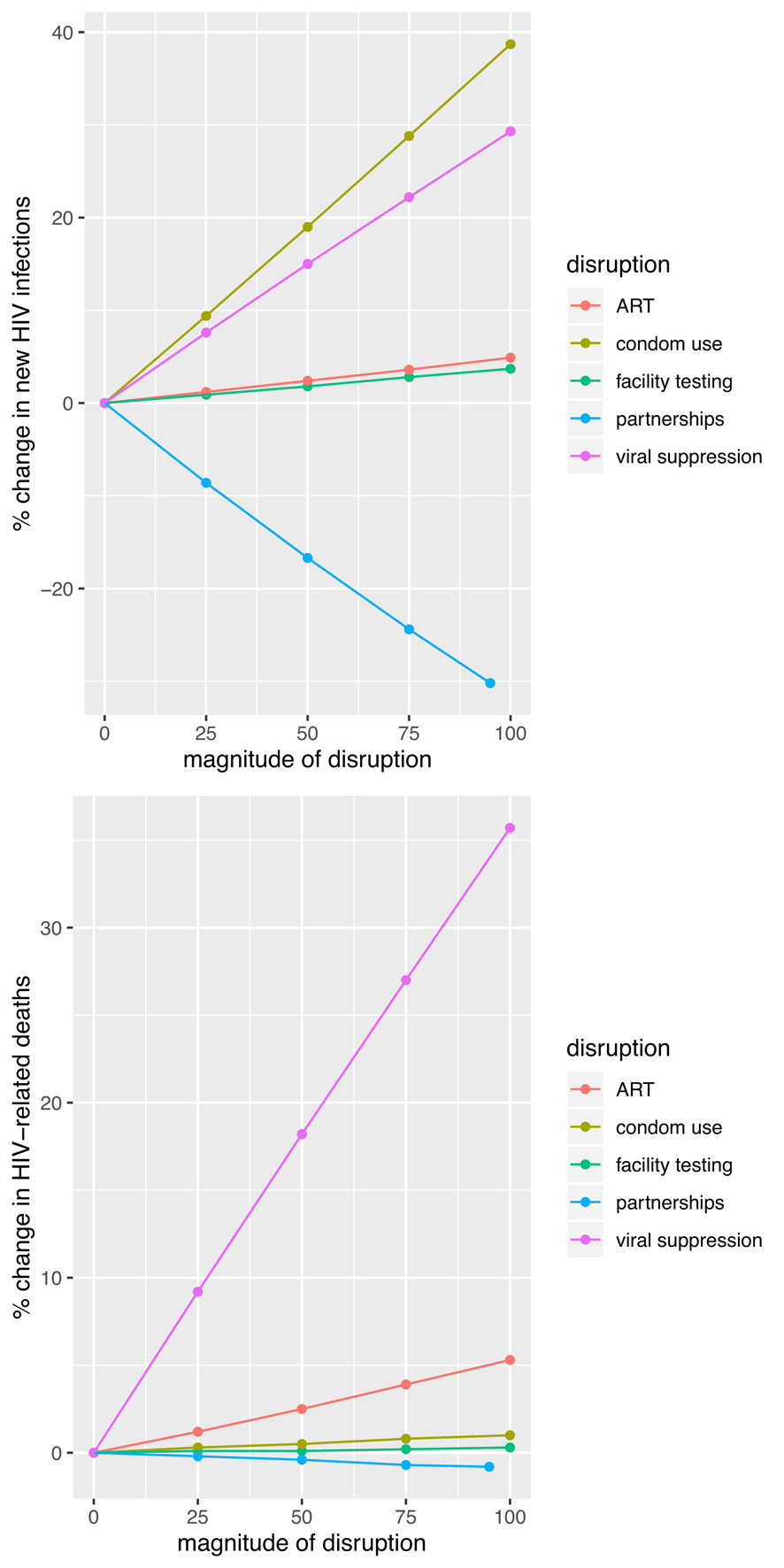

**Figure S4:** Uncertainty in the choice of disruption parameter (0 – 100% of value) plotted with the two impact percentage changes in new infections and deaths over 1 years for a 3-month disruption. Full data can be found in Table S7.

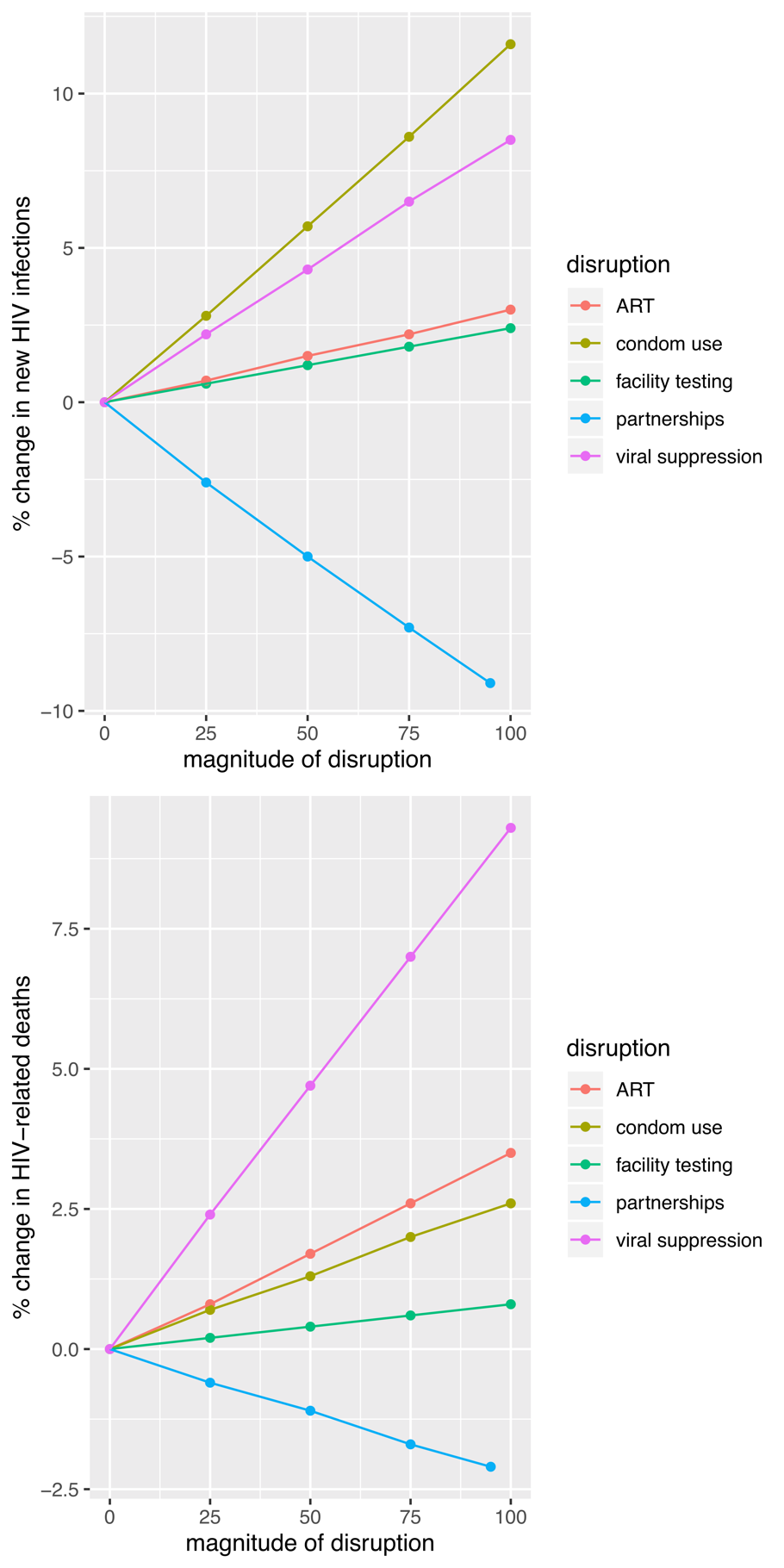

**Figure S5:** Uncertainty in the choice of disruption parameter (0 – 100% of value) plotted with the two impact percentage changes in new infections and deaths over 5 years for a 3-month disruption. Full data can be found in table S8.

| intervention | I1 | %I1 | D1 | %D1 | I1L | I1U | %I1L | %I1U | D1L | D1U | %D1L | %D1U |
| --- | --- | --- | --- | --- | --- | --- | --- | --- | --- | --- | --- | --- |
| A0 | 0 | 0 | 0 | 0 | 0 | 0 | 0 | 0 | 0 | 0 | 0 | 0 |
| A25 | 0.7 | 0.9 | 0 | 0.1 | 0.2 | 5.7 | 0.7 | 1.2 | 0 | 0.2 | 0 | 0.1 |
| A50 | 1.4 | 1.8 | 0.1 | 0.1 | 0.4 | 11.5 | 1.4 | 2.4 | 0 | 0.5 | 0.1 | 0.2 |
| A75 | 2.1 | 2.8 | 0.1 | 0.2 | 0.6 | 17.4 | 2.1 | 3.6 | 0 | 0.7 | 0.1 | 0.3 |
| A100 | 2.8 | 3.7 | 0.1 | 0.3 | 0.8 | 23.4 | 2.8 | 4.8 | 0 | 1 | 0.2 | 0.5 |
| B0 | 0 | 0 | 0 | 0 | 0 | 0 | 0 | 0 | 0 | 0 | 0 | 0 |
| B25 | 0.9 | 1.2 | 0.5 | 1.2 | 0.3 | 6.3 | 0.7 | 1.8 | 0.2 | 3.6 | 1 | 1.4 |
| B50 | 1.9 | 2.4 | 1.1 | 2.5 | 0.5 | 12.9 | 1.5 | 3.6 | 0.4 | 7.4 | 2 | 3 |
| B75 | 2.9 | 3.6 | 1.7 | 3.9 | 0.8 | 19.6 | 2.3 | 5.5 | 0.6 | 11.3 | 3 | 4.5 |
| B100 | 4 | 4.9 | 2.2 | 5.3 | 1.1 | 26.6 | 3.1 | 7.5 | 0.9 | 15.3 | 4 | 6.2 |
| C0 | 0 | 0 | 0 | 0 | 0 | 0 | 0 | 0 | 0 | 0 | 0 | 0 |
| C25 | -6.8 | -8.6 | -0.1 | -0.2 | -48.4 | -1.8 | -10.5 | -7.2 | -0.7 | 0 | -0.4 | -0.1 |
| C50 | -13.2 | -16.7 | -0.2 | -0.4 | -94.3 | -3.5 | -20 | -14.4 | -1.3 | -0.1 | -0.8 | -0.3 |
| C75 | -19 | -24.4 | -0.3 | -0.7 | -137.7 | -5 | -28.5 | -21.4 | -1.9 | -0.1 | -1.2 | -0.4 |
| C95* | -23.4 | -30.2 | -0.4 | -0.8 | -170.6 | -6.2 | -34.8 | -27 | -2.4 | -0.1 | -1.5 | -0.5 |
| D0 | 0 | 0 | 0 | 0 | 0 | 0 | 0 | 0 | 0 | 0 | 0 | 0 |
| D25 | 7.8 | 9.4 | 0.1 | 0.3 | 1.7 | 62.9 | 5.9 | 16 | 0 | 0.8 | 0.1 | 0.5 |
| D50 | 15.8 | 19 | 0.3 | 0.5 | 3.4 | 126 | 11.8 | 32.6 | 0.1 | 1.7 | 0.3 | 1 |
| D75 | 23.8 | 28.8 | 0.4 | 0.8 | 5.2 | 189.6 | 17.6 | 50.6 | 0.1 | 2.6 | 0.4 | 1.6 |
| D100 | 32 | 38.7 | 0.5 | 1 | 7 | 252.5 | 23.4 | 68.5 | 0.1 | 3.5 | 0.5 | 2.2 |
| E0 | 0 | 0 | 0 | 0 | 0 | 0 | 0 | 0 | 0 | 0 | 0 | 0 |
| E25 | 5.7 | 7.6 | 4.3 | 9.2 | 1.6 | 45.8 | 4.4 | 15.4 | 1.4 | 28 | 6.9 | 12.6 |
| E50 | 11.3 | 15 | 8.6 | 18.2 | 3.2 | 90.2 | 8.8 | 30.5 | 2.8 | 55.5 | 13.6 | 25 |
| E75 | 16.9 | 22.2 | 12.8 | 27 | 4.7 | 133.1 | 13 | 45.2 | 4.2 | 82.5 | 20.2 | 37.1 |
| E100 | 22.3 | 29.3 | 16.9 | 35.7 | 6.3 | 174.8 | 17.2 | 59.6 | 5.5 | 109 | 26.8 | 49 |

**Table S7:** Uncertainty in the choice of disruption parameter A-E (0 – 100% of value) given with the total new infections over 1 years (I1) and deaths (D1) and as percentage changes in new infections (%I1) and deaths (%D1). Upper and lower values are given for each e.g. %I1L - %I1U gives the percentage change in new infections over 1 years. Scenarios are as follows: A) Reduction in facility-based HIV testing, B) Reduction in ART initiation, C) Reduction in number of sexual partnerships, D) reduction in condom use, E) Reduction in viral suppression. *C100 is replaced with C95 in order to keep 5% of partnerships (the model requires some partnership information in order to function).

| intervention | I5 | %I5 | D5 | %D5 | I5L | I5U | %I5L | %I5U | D5L | D5U | %D5L | %D5U |
| --- | --- | --- | --- | --- | --- | --- | --- | --- | --- | --- | --- | --- |
| A0 | 0 | 0 | 0 | 0 | 0 | 0 | 0 | 0 | 0 | 0 | 0 | 0 |
| A25 | 1.9 | 0.6 | 0.4 | 0.2 | 0.5 | 17 | 0.4 | 0.8 | 0.1 | 3.1 | 0.1 | 0.3 |
| A50 | 3.9 | 1.2 | 0.7 | 0.4 | 1 | 34.4 | 0.8 | 1.6 | 0.2 | 6.2 | 0.2 | 0.6 |
| A75 | 6 | 1.8 | 1.1 | 0.6 | 1.5 | 52.1 | 1.3 | 2.4 | 0.4 | 9.4 | 0.4 | 0.8 |
| A100 | 8 | 2.4 | 1.5 | 0.8 | 2 | 70.1 | 1.7 | 3.2 | 0.5 | 12.7 | 0.5 | 1.1 |
| B0 | 0 | 0 | 0 | 0 | 0 | 0 | 0 | 0 | 0 | 0 | 0 | 0 |
| B25 | 2.5 | 0.7 | 1.6 | 0.8 | 0.8 | 17.6 | 0.4 | 1.2 | 0.6 | 11.5 | 0.6 | 1 |
| B50 | 5 | 1.5 | 3.3 | 1.7 | 1.6 | 35.7 | 0.8 | 2.5 | 1.2 | 23.4 | 1.3 | 2.1 |
| B75 | 7.7 | 2.2 | 5.1 | 2.6 | 2.5 | 54.2 | 1.2 | 3.8 | 1.8 | 35.7 | 2 | 3.1 |
| B100 | 10.5 | 3 | 6.9 | 3.5 | 3.4 | 73.3 | 1.7 | 5.1 | 2.5 | 48.3 | 2.7 | 4.3 |
| C0 | 0 | 0 | 0 | 0 | 0 | 0 | 0 | 0 | 0 | 0 | 0 | 0 |
| C25 | -9.2 | -2.6 | -1.3 | -0.6 | -59.5 | -2.5 | -3.6 | -1.8 | -8.1 | -0.4 | -1 | -0.4 |
| C50 | -17.6 | -5 | -2.6 | -1.1 | -116.7 | -4.9 | -6.8 | -3.5 | -16.1 | -0.8 | -2 | -0.8 |
| C75 | -25.5 | -7.3 | -3.7 | -1.7 | -171 | -7 | -9.8 | -5.2 | -23.7 | -1.2 | -2.9 | -1.2 |
| C95* | -31.3 | -9.1 | -4.6 | -2.1 | -211.3 | -8.6 | -12.1 | -6.6 | -29.3 | -1.5 | -3.6 | -1.5 |
| D0 | 0 | 0 | 0 | 0 | 0 | 0 | 0 | 0 | 0 | 0 | 0 | 0 |
| D25 | 10.2 | 2.8 | 1.5 | 0.7 | 2.2 | 78.5 | 1.6 | 5.3 | 0.4 | 10.6 | 0.4 | 1.3 |
| D50 | 20.6 | 5.7 | 2.9 | 1.3 | 4.5 | 155.6 | 3.1 | 11 | 0.8 | 21.2 | 0.7 | 2.7 |
| D75 | 31.5 | 8.6 | 4.4 | 2 | 6.7 | 233.1 | 4.7 | 16.8 | 1.2 | 31.5 | 1 | 4.1 |
| D100 | 42.5 | 11.6 | 6 | 2.6 | 9 | 309.9 | 6.3 | 22.8 | 1.6 | 41.7 | 1.4 | 5.6 |
| E0 | 0 | 0 | 0 | 0 | 0 | 42 | 0 | 27.7 | 0 | 0.7 | 0 | 0.8 |
| E25 | 7.4 | 2.2 | 5.1 | 2.4 | 2.1 | 58.2 | 1.1 | 5 | 1.7 | 31.5 | 1.7 | 3.4 |
| E50 | 14.6 | 4.3 | 10 | 4.7 | 4.3 | 114.7 | 2.1 | 9.9 | 3.4 | 62.4 | 3.4 | 6.7 |
| E75 | 21.7 | 6.5 | 14.9 | 7 | 6.3 | 169.6 | 3.2 | 14.6 | 5.1 | 92.7 | 5.1 | 9.9 |
| E100 | 28.6 | 8.5 | 19.6 | 9.3 | 8.4 | 223 | 4.1 | 19.2 | 6.8 | 122.5 | 6.8 | 13.1 |

**Table S8:** Uncertainty in the choice of disruption parameter A-E (0 – 100% of value) given with the total new infections over 5 years (I5) and deaths (D5) and as percentage changes in new infections (%I5) and deaths (%D5). Upper and lower values are given for each e.g. %I5L - %I5U gives the percentage change in new infections over 5 years. Scenarios are as follows: A) Reduction in facility-based HIV testing, B) Reduction in ART initiation, C) Reduction in number of sexual partnerships, D) reduction in condom use, E) Reduction in viral suppression. *C100 is replaced with C95 in order to keep 5% of partnerships (the model requires some partnership information in order to function).
